# Increasing efficacy of contact-tracing applications by user referrals and stricter quarantining

**DOI:** 10.1101/2020.11.30.20240796

**Authors:** Leslie Ann Goldberg, Joost Jorritsma, Júlia Komjáthy, John Lapinskas

## Abstract

We study the effects of two mechanisms which increase the efficacy of contact-tracing applications (CTAs) such as the mobile phone contact-tracing applications that have been used during the COVID-19 epidemic. The first mechanism is the introduction of user referrals. We compare four scenarios for the uptake of CTAs — (1) the *p*% of individuals that use the CTA are chosen randomly, (2) a smaller initial set of randomly-chosen users each refer a contact to use the CTA, achieving *p*% in total, (3) a small initial set of randomly-chosen users each refer around half of their contacts to use the CTA, achieving *p*% in total, and (4) for comparison, an idealised scenario in which the *p*% of the population that uses the CTA is the *p*% with the most contacts. Using agent-based epidemiological models incorporating a geometric space, we find that, even when the uptake percentage *p*% is small, CTAs are an effective tool for mitigating the spread of the epidemic in all scenarios. Moreover, user referrals significantly improve efficacy. In addition, it turns out that user referrals reduce the quarantine load. The second mechanism for increasing the efficacy of CTAs is tuning the severity of quarantine measures. Our modelling shows that using CTAs with mild quarantine measures is effective in reducing the maximum hospital load and the number of people who become ill, but leads to a relatively high quarantine load, which may cause economic disruption. Fortunately, under stricter quarantine measures, the advantages are maintained but the quarantine load is reduced. Our models incorporate geometric inhomogeneous random graphs to study the effects of the presence of super-spreaders and of the absence of long-distant contacts (e.g., through travel restrictions) on our conclusions.

## 1 Introduction

Agent-based epidemiological models allow a population to be embedded in a geometric space to capture the effect of the “local” and “long-distance” contacts that arise in real populations. We use a stochastic susceptible-exposed-infected-removed (SEIR) type agent-based model to study the efficacy of contact-tracing applications such as the mobile phone contact-tracing applications that have been introduced during the COVID-19 epidemic. Our simulation study compares the efficacy of four different uptake scenarios for the contact-tracing application (CTA), combined with the effect of quarantining measures of various strengths. We compare four uptake scenarios (defined in Section 1.3): (i) the users of the CTA are a random subset of the population, the users are selected by one of two different recommender scenarios, in which randomly-chosen initial users recommend the CTA to some of their contacts ((ii) either to just one of them, or (iii) to roughly half of them) who also use the CTA, and finally (iv) a (hypothetical) degree-targetted uptake scenario, in which CTA-users are individuals with the highest number of connections. In each of these CTA-uptake scenarios, we assume that individuals in the population will quarantine themselves as soon as their symptoms are sufficiently strong (exceeding a certain threshold). At this point, users of the CTA also notify their CTA-using contacts, who also quarantine themselves. We examine the combined effect of differing CTA-uptake scenarios and different symptom thresholds for quarantine. We decouple the effects of CTA-uptake scenarios and symptom thresholds from the effects of testing delays, which have been studied in [1]. In order to focus on the former, we work in an idealised hypothetical scenario where testing is immediate upon symptom onset (or where there is only a single virus present in the population that could cause the symptoms) and where the symptom-severity does not influence the probability of a virus transmission between two individuals. We emphasise that our study is qualitative, rather than quantitative, and we aim for universally valid observations in terms of the performance of CTA-uptake scenarios.

### Main observations

In our first experiment (Section 1.3) we qualitatively compare the four uptake scenarios. In all uptake scenarios, we find that contact-tracing applications (CTAs) are effective in decreasing the size of an epidemic (the total number of people who become ill) and in decreasing the maximum number of people who are simultaneously hospitalised, and typically this effectiveness increases linearly or super-linearly with the percentage of the population that use CTA. In these respects, we show that CTAs are effective even with low uptake rates. This confirms the results of a recent study modelling CTAs in Washington state [2]. The novelty of our study is to compare recommendation-based uptake with random uptake, where we find that recommending can significantly improve the efficacy of the CTA. In brief, we find that in scenarios where the CTA is recommended to acquaintances the epidemic size and maximum hospital load decrease at a much higher rate than the rate that would be achieved by randomly selecting the same number of CTA users. One might expect these advantages of recommendation-based uptake to come at the cost of increased quarantine. This is true, however in networks with fewer super-spreaders the quarantine load decreases with uptake percentage (after an initial increase). Once the uptake percentage is sufficiently high, recommendation-based uptake leads to less quarantine, rather than more. Finally, we emphasise that the uptake percentage necessary to completely eradicate the epidemic from the population is generally very high, as was also found in [3], though this is not the focus of this article.

In our second experiment (Section 1.4) we study the impact of the severity of quarantine measures, both on the epidemic itself and on the economic disruption that it may cause (measured in terms of the time that people spend in quarantine over the course of the epidemic). Already under “mild” quarantine measures, the maximum hospital load is drastically reduced and the size of the epidemic is somewhat reduced. However, the economic disruption caused by quarantine is very high. Perhaps surprisingly, imposing stricter quarantine measures (i.e., quarantining individuals already with less severe symptoms), ensures not only that the epidemic is better contained, but also that the economic disruption is lower (under strict enough social distancing measures, travel restrictions, and sufficiently high app uptake). The critical point on the ‘quarantine-strictness scale’ above which the social disruption starts decreasing comes earlier in recommender scenarios and degree-targetted uptake than in random uptake, and even earlier if individuals are more restricted in their movements, i.e., the underlying contact network does not have many long-range connection, ranging over large spatial distances.

As a disclaimer, we recall that we study a case where a *single* SEIR (susceptible-exposed-infected-removed) epidemic is present in the population (see Section 2) and individuals who are not infected do not show any symptoms similar to those that are caused by the virus under investigation. This might be unrealistic for certain types of diseases, but might be a better approximation of reality for others. Finally, we emphasise that we aim for qualitative, comparative results that are robust against changes in the dynamics and the underlying contact network. We refrain from making numerical predictions. *Organisation*. In the remainder of this section we describe the underlying contact network, the CTA-uptake scenarios and varying quarantine measures that we study. We then explain and interpret our main findings regarding each of these. In Sections 2 and 3 we define the agent-based model that we use and the underlying contact networks that we use to model the population.

### 1.1 Networks to model the population

We use a so-called SEIR-type (susceptible-exposed-infected-removed) agent-based models to model the spread of the epidemic, where individuals may or may not show symptoms. The model is defined precisely in Section 2. Roughly, each agent goes through the following stages upon infection: exposed, infected, either symptomatic or asymptomatic, and finally removed. Upon showing symptoms, individuals using the CTA send notifications to the other CTA users that they are in contact with, who then move to quarantine for a fixed duration of time. The model involves a *quarantining* scheme: agents who show sufficiently severe symptoms and agents notified via the CTA both move into quarantine.

Next we briefly discuss the underlying contact network of the agents. We assume that a population is embedded in a geometric space, and the connections between individuals are modelled using a random network model called a Geometric Inhomogenous Random Graph (GIRG) [4]. We define the model in Section 3, while here we only highlight its main features. The GIRG model is a state-of-the-art model for real-world social and technological networks, embedded in a geometric space. We think of the nodes of the network as individuals, each of which has a fixed location in space. The neighbours of a node *u* are nodes with a direct connection (also called a link or an edge) to *u*. A connection may correspond to a friendship or an acquaintance, or simply a contact event, and is correlated with, but does not necessarily coincide with spatial proximity.

There are two robust parameters of GIRGs that control the qualitative features of the network: a parameter *τ* controls the variability in the number of neighbours that individual nodes have (the number of neighbours of a node is called its “degree”). Smaller values of *τ* correspond to more variability, while keeping the average the same. A second parameter *α* controls the number of longrange edges: long-range edges would tend to be present in populations without travel restrictions, (as observed in real-life contact networks, see e.g. [5, 6, 7]).

A value of *α* close to 1 corresponds to having many long-range edges, in this case the network resembles an ageometric (or mean-field) network model (for more on this see Section 3.1 below); while increasing *α* reduces the number of long-range contacts, thus transforming the network into one where the underlying geometry is intrisically more apparent. The following summary will help the reader to understand the experimental results reported below.

- *τ* = 2.3: a GIRG with plenty of “super-spreaders” (nodes with many connections) due to degree variability,
- *τ* = 3.3: a GIRG with fewer super-spreaders,
- *α* = 1.3: a GIRG with many long-range edges, hence the underlying geometry is “less apparent”
- *α* = 2.3 a “more geometric” GIRG, i.e., a network that resembles a lattice better, since it does not have too many long-range connections.

The parameter values (*τ, α*) are chosen to represent *each universality class* of GIRGs with respect to average distance in the network. See more details and explanation in Section 3.

For comparison, we also used a common uncorrelated network model, the *configuration model*, corresponding to a network with no underlying geometry, see details in Section 3.2. Configuration models do not have an associated *α* parameter expressing the strength of geometry, though there is still a parameter *τ* indicating the presence of many (or fewer) super-spreaders. To make comparison to GIRGs possible, also in the configuration model we take *τ* = 2.3 for modelling a network with plenty of superspreaders and *τ* = 3.3 for a model with fewer superspreaders, that again correspond to the universality classes of these models with respect to average distance. We tune the parameters of all underlying networks so that they have the same average degree and size.

Another family of commonly-used correlated network models are *spatial preferential attachment* models [8, 9]. GIRGs have two key advantages over these models for our purposes. The first is that they are much simpler to analyse from a theoretical perspective due to the large amount of independence present in the model. The second is that long-range edges are key to our paper as a way of modelling travel restrictions. While some spatial preferential attachment models do allow long-range edges [10], these were not the original focus of the model and there is no universally-agreed standard; by contrast, the *α* parameter of GIRGs fits our needs well.

### 1.2 Key performance indicators (KPIs)

To compare the performance of the four uptake scenarios as well as the strictness of quarantining, we study three KPIs as a function of the parameters.

- Size: The size of the epidemic, meaning the total number of individuals ever infected during the whole course of the epidemic.
- HMax: The (approximated) maximum hospital load. In our study, we define the hospital load at any time to be 5% of the number of agents in infected states (symptomatic or asymptomatic) individuals. Of course, the 5% is just a scaling, and changing this factor does not change the *shape* of the curve. HMax is then the maximum of this hospital load, over the course of the epidemic.
- Quar: The average number of days spend in quarantine per person, over the course of the epidemic. We obtain this quantity as the accumulated number of days spent in quarantine by the population, divided by the total population size.

### 1.3 Experiment 1: Application-uptake scenarios

An important aspect of our study is a comparison of the efficacy of four different CTA-uptake scenarios. Suppose that *p*% of the population use and also comply with a CTA, in the sense of reporting symptoms and also going into self-quarantine when instructed to do so. We compare the effectiveness of the CTA in terms of reducing the total size of the epidemic (which we denote Size) and the maximum hospital load HMax, when this *p*% uptake is achieved in four different ways:

1. randomly chosen: *p*% of the population uses the CTA, and this *p*% is chosen uniformly at random,
2. via basic recommendation: a set of “initial users” is chosen uniformly at random. Each initial user recommends the CTA to a single uniformly-chosen neighbour who then also uses it,
3. via ring recommendation: each initial user recommends the CTA to its neighbours, each of them will use the app with probability 1*/*2 independently of the rest,
4. via degree targetting: the *p*% of the population with the highest number of connections use the CTA.

In terms of controlling the epidemic, one would expect that it is desirable if the *p*% most influential members of the population use the CTA, rather than a randomly-chosen *p*%. However, *targetting* the application in this way incurs cost (it is necessary to determine which members of the population have the most influence, and to persuade them to use the application). The recommender scenarios provide a less-costly, easier-to-implement solution. Each time a (randomly-chosen) member of the population begins using the CTA, this user is asked to persuade either a single, randomly-chosen contact or all of its contacts to also use the CTA. Informally, these contacts are friends or acquaintances. In our network model, they correspond to neighbours in the network. See Section 1.1 and Definition 4 for details. In the case where all contacts are asked to use the CTA, our model assumes that roughly half of these contacts do begin using the CTA. This ring recommendation method has some connections with a certain vaccination protocol, called ring vaccination, hence the name. Ring vaccination has been successful in the past to eradicate small pox [11] and has also been used against Ebola [12]. Such recommender scenarios have previously been shown to be effective in the context of vaccines [13] on synthetic models. Note that, to achieve the same uptake percentage *p*%, the initial percentage of users for the recommender-based scenarios is *smaller* than *p*%, see Fig 1.

**Figure 1.**
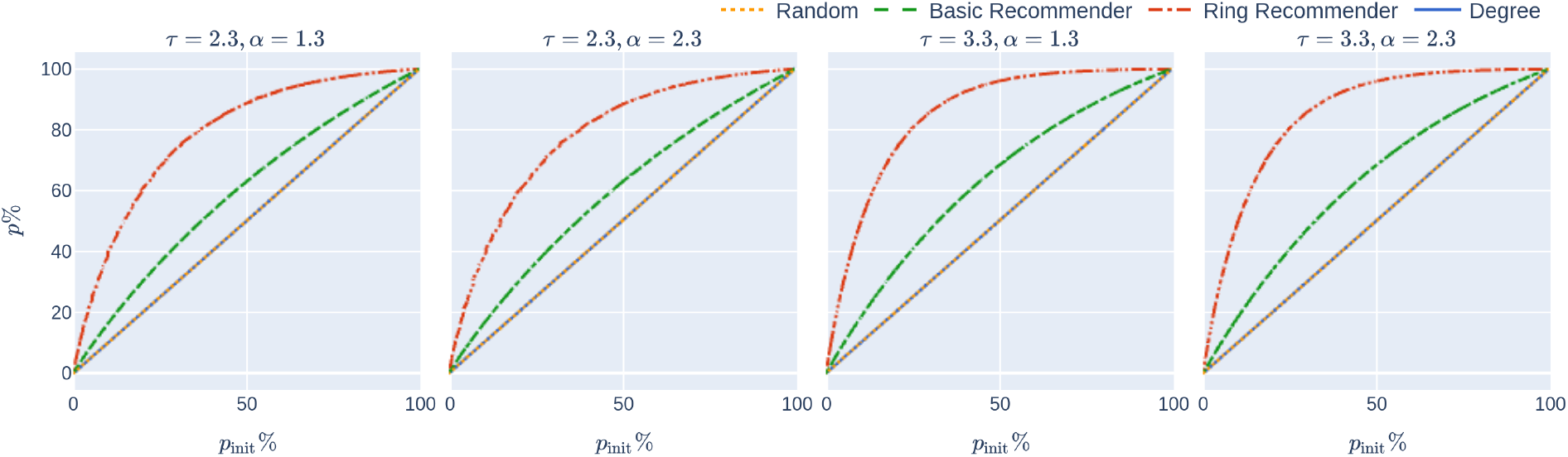
: The initial app uptake *p*_init_% in relation to the final app uptake *p*% on four networks and the different uptake scenarios. The curves for random uptake degree-targetted uptake coincide and are straight lines, since it holds that *p*% = *p*_init_%, while for recommender scenarios *p*% *> p*_init_%. Increasing *p*% increases the probability that a node is recommended by multiple persons, explaining the concave curves corresponding to the recommender scenarios.

Basic recommendation also has connections to a vaccination protocol called acquaintance vaccination which typically outperforms random vaccination in network models [14, 15]. The main difference between the two scenarios is that in acquaintance vaccination, only the neighbours of the randomly-chosen set of “initial users” receive the vaccine rather than the users themselves. This makes sense in the context of trying to allocate a limited supply of vaccines, but not in our context — it is reasonable to assume that anyone recommending the app to a friend will also install it themselves.

## Results

We varied the CTA-uptake percentage for the four uptake scenarios on six networks, each with 500, 000 individuals and average degree 13 (average degree 13 roughly corresponds to empirical findings concerning the number of contacts per individual [16]), using the network models from Section 1.1. In each case, we studied the effect of the CTA-uptake percentage and the uptake scenario (randomly chosen, degree-targetted, basic recommender or ring recommender) on the three KPIs from Section 1.2. We explain our results using the GIRGs from Section 1.1 with *τ* ∈ {2.3, 3.3} and *α* = {1.3, 2.3}. The last two networks mentioned in Section 1.1, modelled by the configuration model, are merely for comparison and are included in our later figures.

### 1.3.1 Hospital load and Epidemic Size: Recommenders perform very well, ring recommender best

The main message indicated by our simulations (row 2 of Fig 2) is that introducing recommender scenarios strongly increases the efficacy of a CTA in terms of reducing hospital load: Having achieved uptake *p*% via any recommender scenario is significantly more desirable (in terms of reducing the maximum hospital load) than having achieved *p*% uptake by randomly chosen members of the population. Furthermore, the degree-targetted scenario works best, and the ring-recommender scenario is almost as good as this. Adding the subscripts rand, deg, basic, and ring to indicate the uptake scenarios, we find typically that

**Figure 2.**
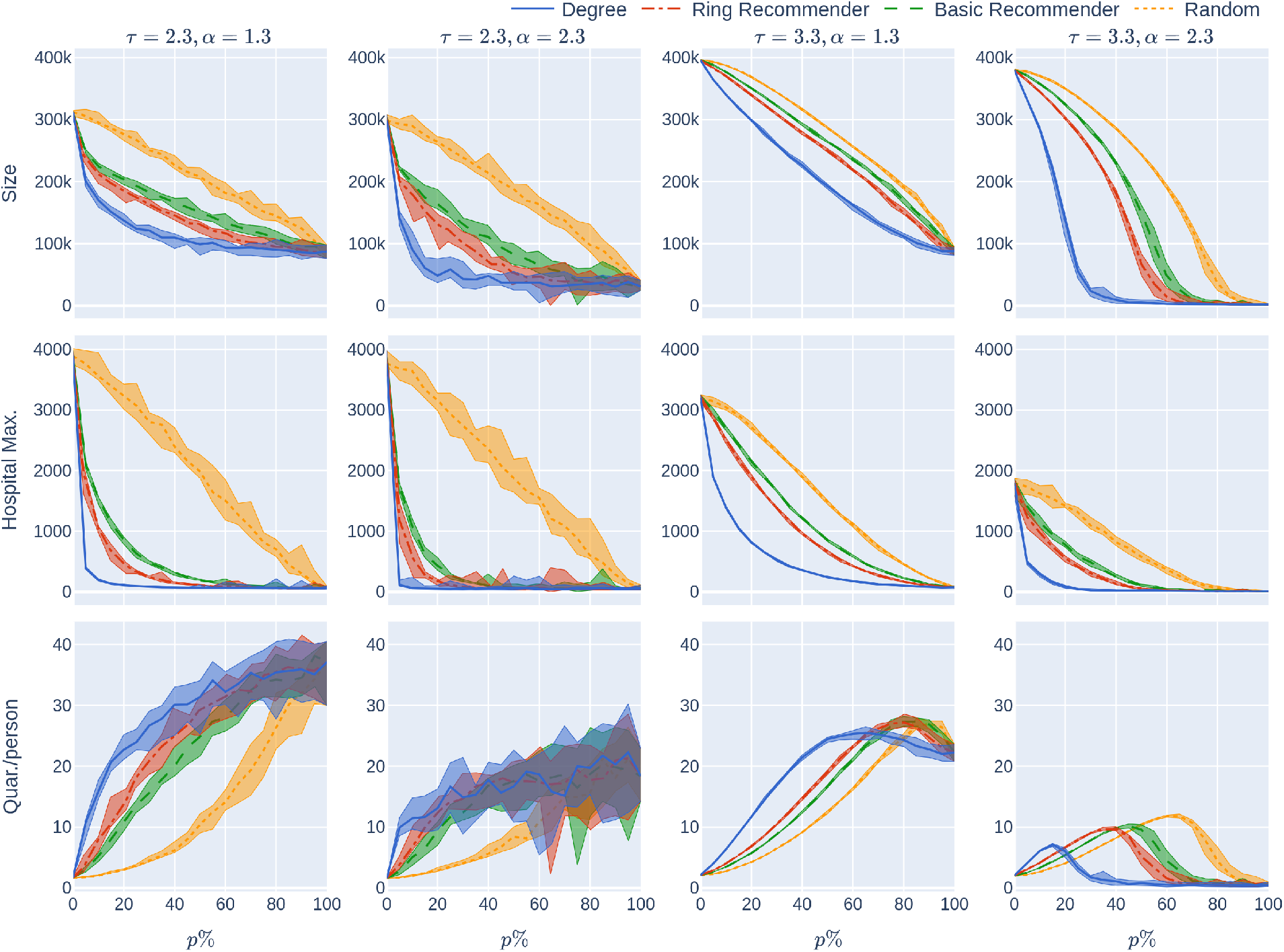
: The KPIs (Size, HMax, Quar) as a function of the CTA-uptake percentage *p*% for the four CTA-uptake scenarios can be found in the rows. The population is modelled using four 500,000 node GIRG networks where *τ* = 2.3 has more super-spreaders and *τ* = 3.3 has fewer super-spreaders, *α* = 1.3 is less geometric and *α* = 2.3 is more geometric, arranged in the four columns. The *x*-axis shows the uptake percentage *p*% varying from 0 to 100% at step size 5%. Simulations are done for the 21 values of *p*% corresponding to these steps. The *y*-axis shows the corresponding value of the KPI. The four uptake scenarios correspond to the four curves on each figure. For each parameter value, the plotted result is the median over 10 runs. The shaded region around the plot covers the results of all 10 simulations. The epidemic is started by infecting 100 individuals, chosen uniformly at random. Simulation is halted when there are no more exposed or infected vertices. For completeness (the reader does not need to know this at this point, but it will be useful for comparison later) the simulations use *β* = 0.05 and *q* = 0.6. *β* is the rate of infection. It is defined in Section 2, and this value corresponds to a low rate of infection, for example due to social distancing measures. The parameter *q* denotes the severity of the quarantine measures. It is studied later, in Experiment 2.

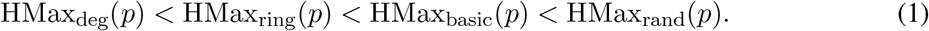

It is worth explaining at this point what we mean by “typically” and in what sense Equation (1) is intended. Obviously, it is not strictly true — for example if the uptake percentage *p*% is 0% or 100%, then all uptake scenarios are equivalent. Furthermore, our results are numerical, so will have some inaccuracies. As can be seen from the 2nd column of row 2 of the figure, if the CTA works well enough to suppress the epidemic then the various uptake scenarios are roughly equivalent, and cannot be compared. What we mean by “typically” is that, as is apparent from row 2, when the CTA uptake does matter, our data strongly suggests that the hospital max is smallest under the degree-based scenario, next smallest under the ring scenario, and largest (by some measure!) under the random scenario.

The same inequalities typically hold true for the size of the epidemic (first row of Fig 2):

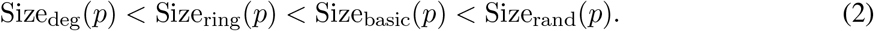

For a different depiction of this phenomenon at *p* = 5% see Fig 3, where the course of the epidemic is plotted against *time*. The epidemic curve on the left represents the total number of infected individuals (during the course of the epidemic). The epidemic curve on the right represents the number of individuals that are currently infected. The figure demonstrates how the random-uptake scenario is outperformed by the other uptake scenarios.

**Figure 3.**
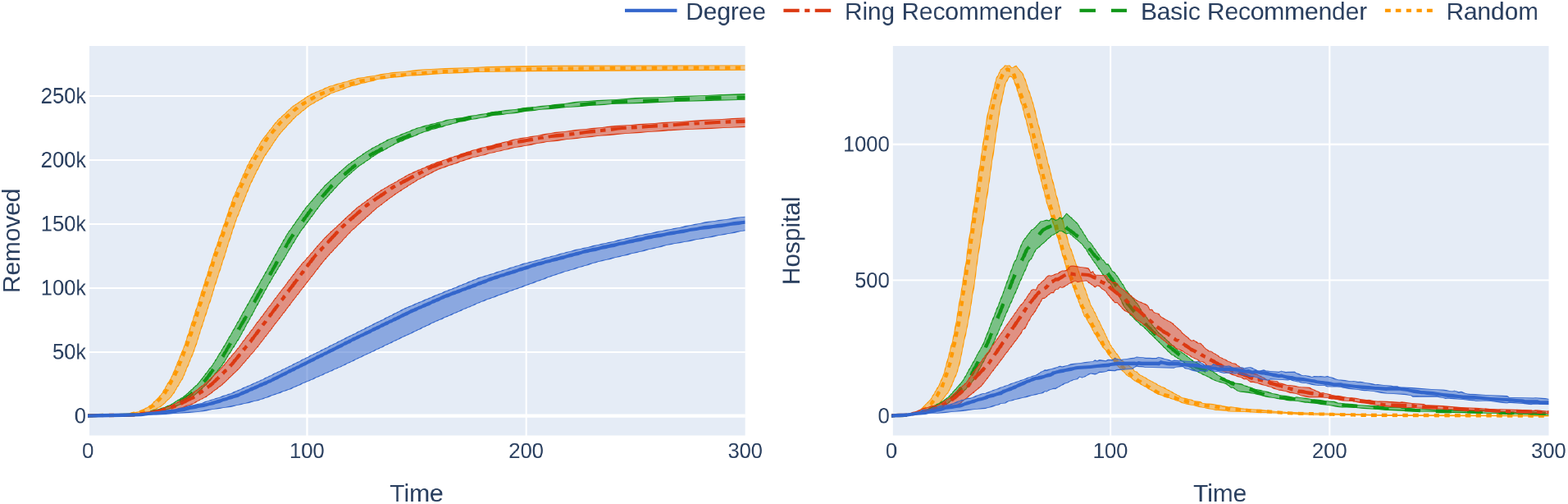
: Simulations of an epidemic on a 500,000 node GIRG network with few super-spreaders (*τ* = 3.3) and less geometry (*α* = 1.3), with uptake percentage *p*% = 55%. The *x*-axis shows the time steps over which the epidemic is simulated. The *y*-axis of the left figure shows the number of removed individuals over the course of the epidemic. The *y*-axis of the right figure shows the number of hospitalised individuals over time (in the same simulation). The plotted curves correspond to the median over 20 runs (in the four scenarios); the shaded area covers the 25th-75th percentile of these runs over time. The left figure shows that the random uptake scenario is outperformed by all other scenarios and has a steeper incline, leading to a situation where more people get infected. The right figure shows that the number of hospitalised individuals is also higher under the random uptake scenario. This figure corresponds to the *p* = 55% value in the first two rows of the leftmost column of Figure 2. Similarly to the situation regarding that figure, the epidemic is started by infecting 100 individuals, chosen uniformly at random. Simulation is halted when there are no more exposed or infected vertices. For completeness the simulations use *β* = 0.05 and *q* = 0.6.

The reason that recommender scenarios always perform better than the random uptake scenario is explained by the so-called friendship-paradox [17]: a friend of a random user is likely to have a higher number of contacts than the random user itself, so a recommender scenario intrinsically finds users with high numbers of connections. This effect is more pronounced when the node-degrees are highly varying (*τ* = 2.3) and it is even more pronounced in the ring-recommendation scenario. The reason for this is that ring recommendation introduces highly connected clusters of CTA users, and it is hard for the epidemic to thrive in such an environment: the CTA clusters block its spreading.

### 1.3.2 Improving CTA efficacy by slightly increasing uptake rates

We study the shape of the curves Size_□_(*p*) and HMax_□_(*p*) as *p* varies and □ = rand, basic, ring, deg. Intuitively, we examine what happens when we slightly increase the CTA-uptake percentage, especially in the (at least in Europe) more realistic low-uptake regimes (*<* 50%).

Our first finding (see the second row of Fig 2) is that random uptake decreases HMax roughly *linearly* in the uptake percentage *p*%. Here we don’t mean that the plotted yellow curves in the second row (which are numerical approximations to Hmax_rand_(*p*)) are precisely linear — instead we mean that the rate at which the hospital maximum decreases is *roughly* constant, as the uptake percentage *p*% increases. This rate of decrease depends on the underlying network (the four columns).

It also depends on the rate of the infection, *β*, which is fixed in this section. In our experiments we later vary *β* (see Fig 8). For higher values of *β* it appears that HMax decreases concavely as the uptake percentage *p*% increases, meaning that HMax decreases more quickly for larger percentages *p*%.

A similar phenomenon holds for the epidemic size Size_rand_(*p*) (see the first row of Fig 2) when *τ* = 2.3 so there are many super-spreaders in the network, while the curve is more concave when *τ* = 3.3 (fewer super-spreaders). See also Fig 9, where the infection rate *β* is varied. Note that Size(0) and Hmax(0) do not depend on the uptake scenario, since no individual is using the CTA when *p* = 0.

We explain why the approximately linear decline is intuitively surprising, where it appears. For an infection to be prevented via the CTA, both the infector and the infected individual have to use the CTA, and the chance of this by random uptake is (*p/*100)^2^ ≪ *p/*100. Thus one’s first guess would be that HMax_rand_(*p*) and Size_rand_(*p*) would be concave in *p*, not linear, over all infectiousness parameters. Yet, we have sees that the curve is concave only in scenarios with high infectiousness and in contact networks with strong underlying geometry (see Figs 8 and 9, where the infectiousness parameter (*β*) is varied). The conclusion is that, even with random uptake, the CTA works better than might be expected, decreasing Size and HMax roughly linearly for a large range of parameter values (of infectiousness, and of the underlying network), rather than concavely.

The recommender scenarios react even better to small increases in the uptake percentage (for small values of *p*). Our experiments show (see the first row of Figure 2) that all recommender scenarios (and especially the ring-recommender scenario) react more strongly (than the random-uptake scenario) to a slight increase in the uptake percentage *p*%. That is, the epidemic size curves decrease at a faster rate under basic and ring recommendation, as compared to under the random uptake scenario. Roughly, we find that for *p*% *<* 50% and a small positive number d*p*,

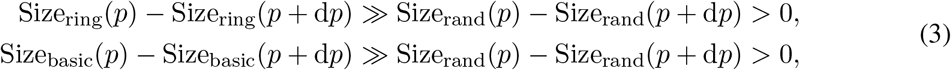

meaning that (apart from perhaps for *τ* = 3.3, *α* = 1.3), the total size of the epidemic as a function of *p* decreases substantially more steeply for ring-recommender and basic-recommender uptake than for the random uptake. While this is true for both *τ* = 2.3 and *τ* = 3.3, in the situation where there are many super-spreaders (*τ* = 2.3), the curves seem in addition to be convex (meaning that the rate of decay is highest when the uptake percentage is small).

For the maximum hospital load HMax, the effect is similar, but stronger (see row 2 of Figure 2). Here the HMax curve is a convex curve for all recommender scenarios.

In summary, we find that for many underlying contact networks, the functions Size_ring_(*p*), Size_basic_(*p*), HMax_ring_(*p*), and HMax_basic_(*p*) are convex, having a steep decline when *p*% *<* 50%, while Size_rand_(*p*) and HMax_rand_(*p*) are linear or concave functions. This is illustrated further in Figures 8 and 9 where the level of infectiousness is varied: the phenomenon is robust in the “supercritical” regime where there is a large outbreak, apart from in graphs with few super-spreaders in situations with very high infection rates. In this latter case the curves might coincide or are more concave.

### 1.3.3 Geometry helps the CTA

A less obvious observation that we make is that (considering networks with the same average number of connections per node), the more geometric the underlying contact network is, the better the performance of the CTA in terms of reducing the size of the epidemic and the maximum hospital load, across the four uptake scenarios. In other words, lack of long-range connections helps CTAs. This agrees with existing work on classical contact tracing [18, 19], despite the increased heterogeneity introduced by random app uptake; see [20] for a detailed survey.

Moreover, the positive effect of recommending becomes more exaggerated in geometric networks, see Fig 4. Emphasising in notation by adding a superscript ‘geo’ and ‘ageo’ to emphasise whether the underlying network has strong geometry (*α* = 2.3) or less geometry (*α* = 1.3), we typically find that (for fixed *τ* and reasonable uptake percentages)

**Figure 4.**
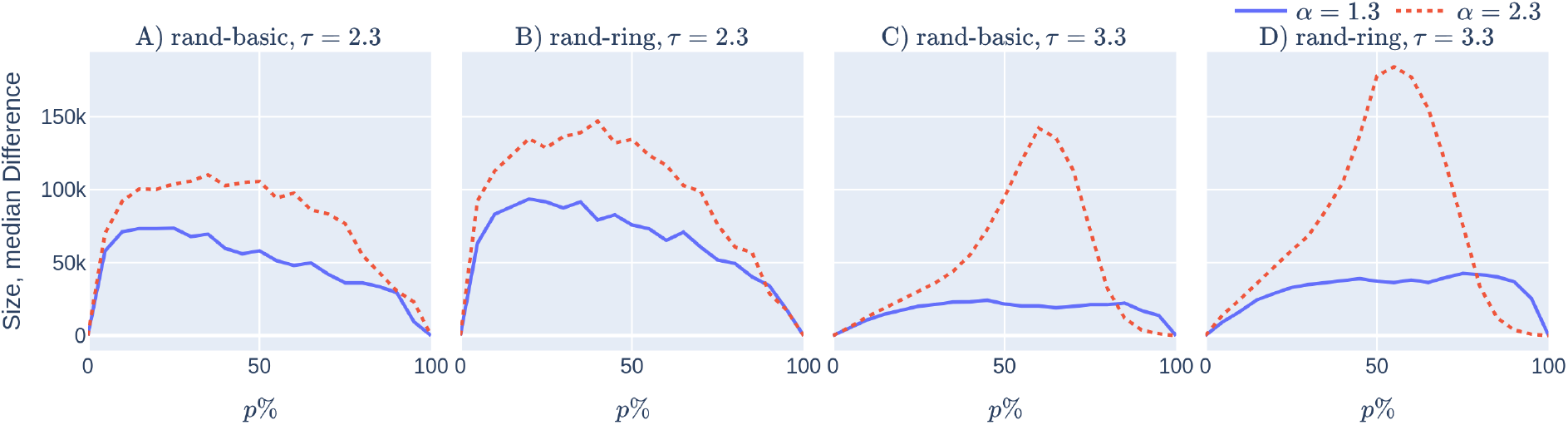
: The reduction in epidemic size achieved by CTAs with user recommendation improves when the network has stronger geometry (for all values of *p*% *<* 85%). Subfigures (A) and (C): The red dashed curve shows 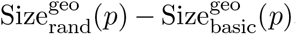, while the blue continuous curve shows 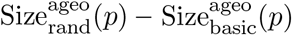 as a function of the uptake percentage. In subfigures (B), and (D), the red dashed curve shows 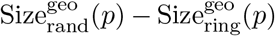, while the blue continuous curve shows 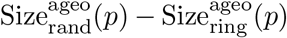as a function of the uptake percentage *p*%. Observe that the red dashed curves are (typically) above the blue curves. In subfigures (A) and (B) the underlying network contains many super-spreaders, while in subfigures (C) and (D) the network contains fewer: observe that the presence of fewer super-spreaders makes the red dashed curve, the epidemic size difference in geometric networks between recommender and random uptake, more spiky. The simulation details are the same as those in Figure 2. The same four 500,000 nodes GIRGs are used. The *x*-axis shows the uptake percentage *p*% varying from 0 to 100% at step size 5%. Simulations are done for the 21 values of *p*% corresponding to these steps. For each parameter value, the plotted result is the median over 10 runs. The epidemic is started by infecting 100 individuals, chosen uniformly at random. Simulation is halted when there are no more exposed or infected vertices. The simulations use *β* = 0.05 and *q* = 0.6.

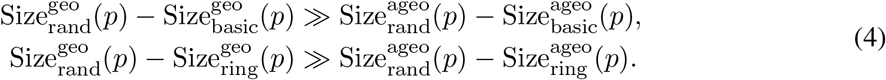

This is illustrated on Fig 4, where the epidemic size difference (corresponding to the number of infections that are prevented by the recommendation) is plotted. Observe that the dashed red curve, corresponding to the epidemic size difference in geometric networks stays above the blue curve, corresponding to the epidemic size difference in ageometric networks. We emphasise that both curves are positive, i.e., recommendation performs better than random uptake in both geometric and ageometric networks.

The intuitive explanation for Equation (4) is the same as what we gave in Section 1.3.1: recommending has two main effects. The first effect is finding the super-spreaders via the friendship-paradox effect. The second effect of recommending is only present in geometric networks: recommending also forms geometric barriers of CTA-users around a central node. These barriers are hard for the infection to pass through.

On the empirical side, we mention that a way to introduce more geometry into the underlying contact network is via travel restrictions. Two of the authors previously studied the effect of such intervention on epidemic curves in [21], which found that already in simple scenarios where there are no CTAs, travel restrictions are indeed most effective in reducing the height of the (first) peak.

### 1.3.4 How costly is quarantine?

A common belief is that using a CTA might cause a high quarantine load on the population. However, row 3 of Fig 2 demonstrates that, at least in the case of fewer super-spreaders (*τ* = 3.3) the average number of days that an individual has to quarantine does not rise very steeply with the CTA-uptake percentage *p*%. Note that the date for the case with more superspreaders (*τ* = 2.3) has very high variance, so we do not draw conclusions abou this case. Note also that the conclusions are relevant in situations where social-distancing measures make the underlying infection rate not too high, this corresponds to the choice of *β* = 0.05 here. Though it is beyond the scope of this study, note that quarantine time can be reduced by providing prompt testing to people who are notified to quarantine by the CTA.

Here we note a surprising feature that applies to underlying networks with relatively few super-spreaders (*τ* = 3.3) in Fig 2. In this case, Quar(*p*) is roughly unimodal, meaning that when the uptake percentage *p*% is small, the amount of quarantine time per person increases as *p* increases. However, roughly, the amount of quarantine is maximised for some *p*^*⋆*^ % *<* 100 %, and decreases above *p*^*⋆*^.

This means that using CTAs only increases the average quarantine time below uptake *p*^*⋆*^ but not above it. We mention that *p*^*⋆*^ *depends on the uptake scenario*, and comes earlier with recommendation scenarios than with random uptake. On the last row of Fig 10 the same phenomenon is observed for various levels of infectiousness of the disease: this rough unimodality in Quar(*p*) is robust for these underlying networks. It disappears, however, for networks where either there are more super-spreaders (so the data is too noisy to draw conclusions) or the geometry is less apparent (Fig 10).

The conclusion is that, in this particular setting, using CTAs at high uptake rates is effective not only for reducing the epidemic size and maximum hospital load of the epidemic but also in reducing the average quarantine length.

### 1.3.5 Other parameter values

The experiments in Fig 2 assumed an epidemic model (see Section 2) with a low rate of infection *β* = 0.05. This means that each infectious node infects each of its susceptible neighbours with probability *β* at every time step (e.g., every day). Low rates of infection can be achieved by social-distancing measures. The results that we have mentioned so far are robust over other infection rates. See Figs 8, 9, and 10 where the rate of infection is varied, for maximum hospital load, epidemic size, and quarantine load.

### 1.4 Experiment 2: Varying the strictness of quarantine measures

It has been observed that the severity of symptoms varies with the individual [22, 23, 24, 25]. In our model, we assume that there is an underlying probability distribution that determines whether an individual would be symptomatic, in the case that this individual is infected, and also that there is an underlying probability distribution determining the severity of these symptoms. Policy makers may then impose quarantine or home isolation on individuals that have symptoms above a certain severity-threshold; setting the threshold is a political/economic decision. In our model, the strictness of quarantine measures is represented by a value *q* ∈ [0, 1]. This quantity *q* is the fraction of individuals whose symptoms would be so severe that they would go into home-quarantine if infected. Strict quarantine measures correspond to large values of *q* – in the extreme, taking *q* = 1 means that all infected individuals would go into home-quarantine. Recall that we work under the assumption that either a single virus is present in the population (so all relevant symptoms are caused by this virus) or testing is immediate and available to those who exceed the symptom severity threshold. This assumption is important for the results in this section.

For any given epidemic, there is an underlying probability *q*_symp_, which is the proportion of infected individuals that experiences any symptoms upon infection (mathematically, the probability of showing symptoms upon infection). Implementing a quarantine-strictness *q > q*_symp_ would require a program of mass testing. The implementation of such a testing program is beyond the scope of this paper, however we make two remarks:

- For many epidemics, the value of *q*_symp_ is fairly high (for COVID-19, the literature is varied, [26] finds a *q*_symp_ ≈0.3 over all population, varying in age, [27, 28] find it ≈0.6 and ≈0.7, respectively, while [29] finds it as high as ≈ 0.85).
- our results indicate that setting quarantine-strictness even below *q*_symp_ can still be effective for decreasing the epidemic size and maximum hospital load.

The social benefits of quarantining measures are nuanced. While quarantining clearly helps to reduce the size of the epidemic (Size) and the maximum hospital load (HMax), it also comes at the high societal cost of reduced work capacity. As a policy maker, one might like to balance the reduction in Size and HMax against Quar, the average number of days spent in quarantine per person, over the course of the epidemic. In Experiment 1 we took the fixed value *q* = 0.6. In Experiment 2, we study how the setting of the quarantine-strictness *q* affects all of these KPIs. In this experiment, we vary the quarantining strength *q* and the CTA-uptake percentage *p*% for the four CTA-uptake scenarios. Our results are summarised in Fig 5, that we elaborate below.

**Figure 5.**
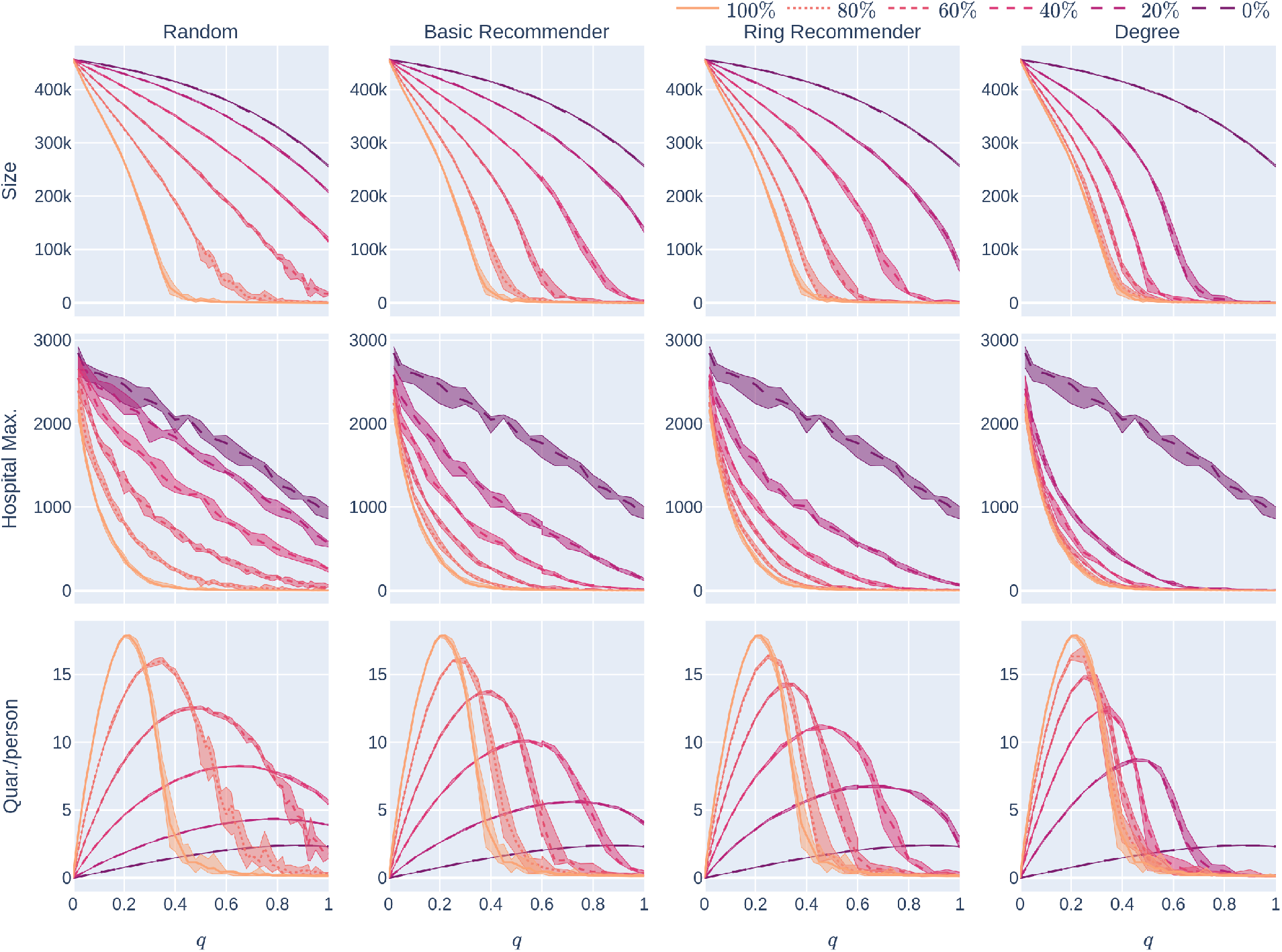
: Influence of the quarantine strictness *q* for various uptake percentages *p*% and the four uptake scenarios: the KPIs are plotted against *q*, the quarantine strictness. We work here with a fixed underlying network, a 500,000 node GIRG with *τ* = 3.3 (fewer super-spreaders) and *α* = 2.3 (more geometric). Each column represents a given uptake scenario (random, basic-recommender and ring-recommender, and degree-targetted). The *x*-axis shows the quarantine strictness *q* varying from 0 to 1 at step size 0.05. Simulations are done for the 21 values of *q* corresponding to these steps. There is an additional simulation point at 0.02 (in order to avoid division by 0 in the computation for HMax and still have a point close to 0). The *y*-axis shows the corresponding value of the KPI. The curves on each figure correspond to the different uptake percentages, as labelled. For each parameter value, the plotted result is the median over 5 runs. The shaded region around the plot covers the results of all 5 simulations. The epidemic is started by infecting 100 individuals, chosen uniformly at random. Simulation is halted when there are no more exposed or infected vertices. The infection rate is *β* = 0.05.

**Figure 6.**
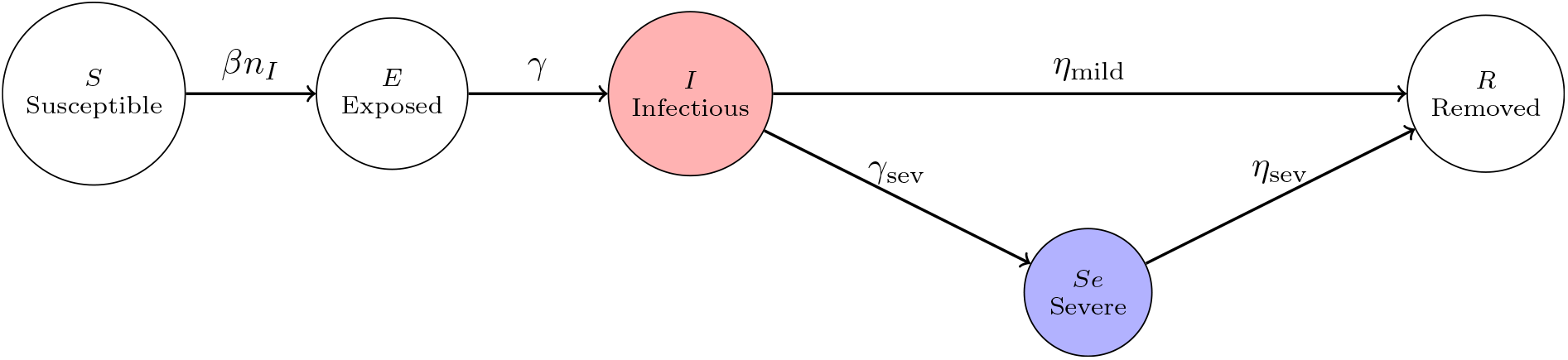
: An illustration of the dynamical change of states. On the first transition arrow, *n*_*I*_ denotes the number of neighbours in state (I) of a susceptible node. Only nodes in 𝒱_sev_ transition to state (Se). By setting 1*/η*_mild_ = 1*/γ*_sev_ + 1*/η*_sev_ the average duration of having the virus in an individual’s system is 1*/η*_mild_ for each node. Only nodes in the red circle can infect, while nodes in the blue circle stay in home isolation.

**Figure 7.**
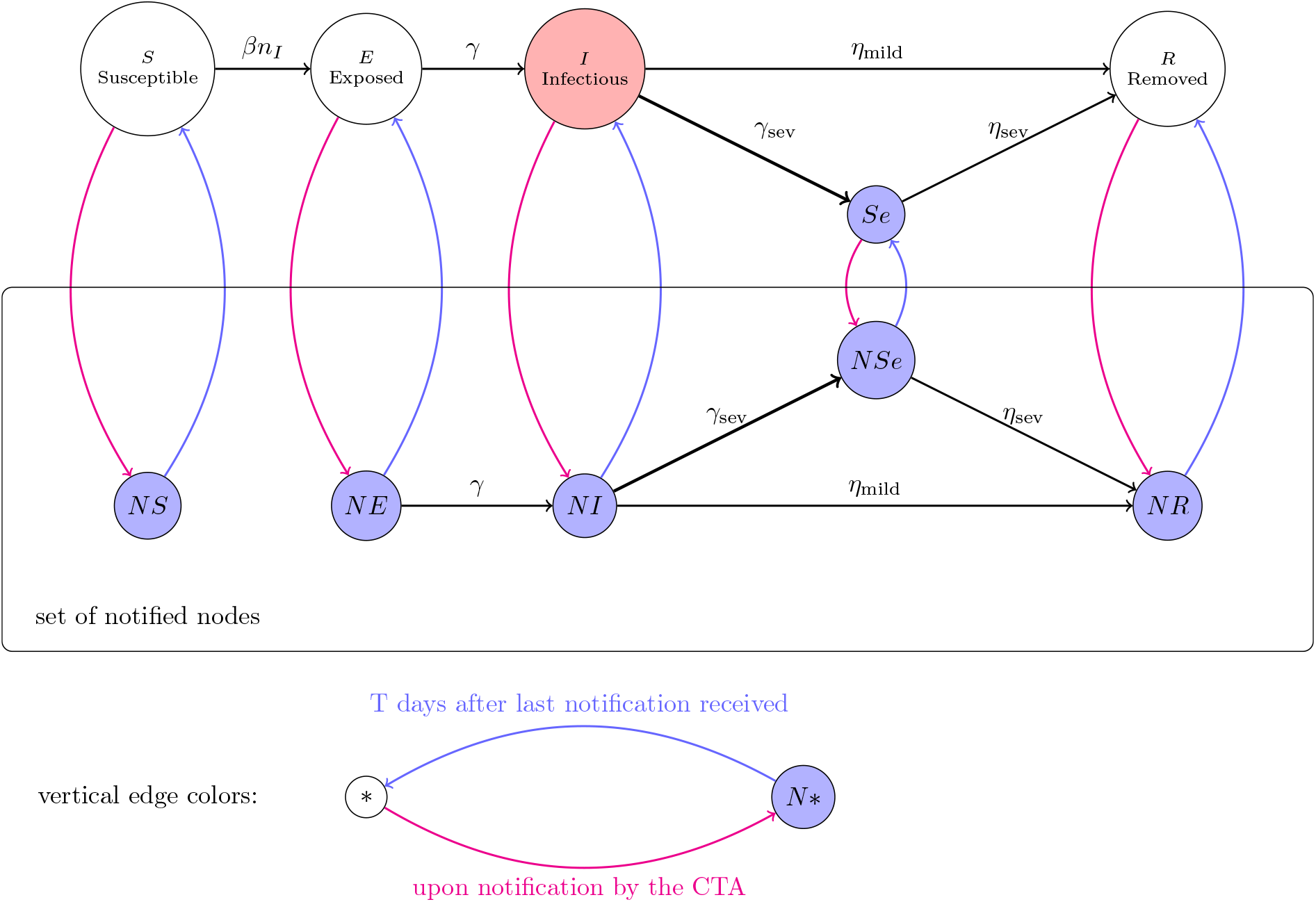
: An illustration of the dynamical state changes in the presence of a contract-tracing application. The black arrows represent the transitions of the SEISeR model and the corresponding transitions in the notified versions of the states. After each time step, nodes move between the *O*-states (S), (E), (I), (Se) and (R) and the *N* -states (NS), (NE), (NI), (NSe) and (NR). Moves to *N* -states (depicted by red arrows) follow from CTA notifications. CTA users (nodes in 𝒱_*u*_) receive such notifications when neighbouring CTA users show sufficient symptoms, thus transitioning from (I) to (Se) or from (NI) to (NSe). Nodes in blue states are required to self-quarantine so cannot spread infection. Moves to *O*-states (depicted by blue arrows) occur when a node has received no new notifications for *T* = 14 days.

**Figure 8.**
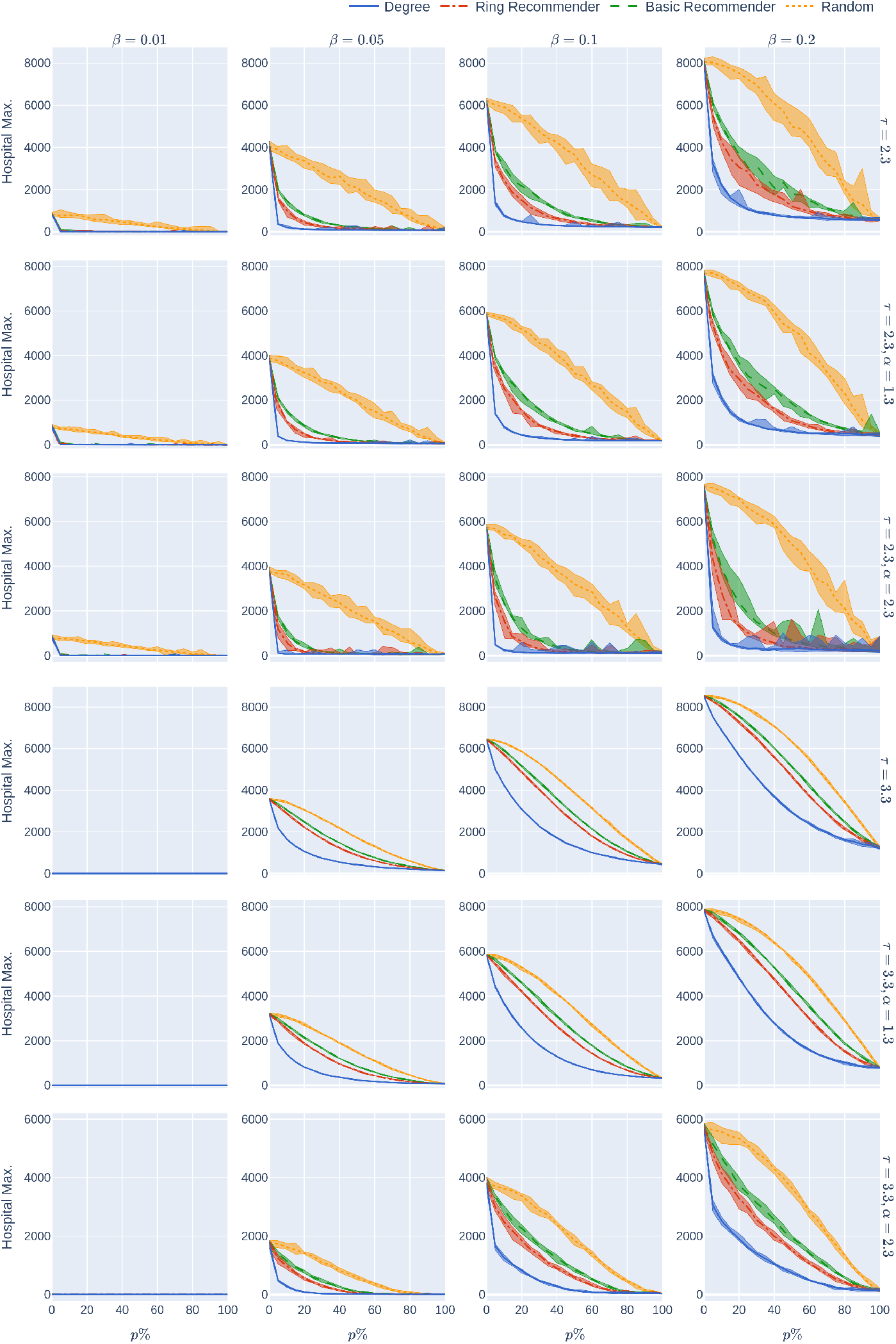
: The maximum hospital load as a function of the uptake percentage *p*%. The population is modelled using four 500,000-node GIRG networks where *τ* = 2.3 has more super-spreaders and *τ* = 3.3 has fewer super-spreaders, *α* = 1.3 is less geometric and *α* = 2.3 is more geometric, arranged in the four rows. For comparison, there are two rows for 500,000-node configuration models with *τ* = 2.3 and *τ* = 3.3. The *x*-axis shows the uptake percentage *p*% varying from 0 to 100% at step size 5%. Simulations are done for the 21 values of *p*% corresponding to these steps. The *y*-axis shows the corresponding value HMax(*p*). Different columns indicate varying infection probabilities *β*. The HMax data in Fig 2 corresponds to the *β* = 0.05 column. As in Figure 2, The four uptake scenarios correspond to the four curves on each figure. For each parameter value, the plotted result is the median over 10 runs. The shaded region around the plot covers the results of all 10 simulations. The epidemic is started by infecting 100 individuals, chosen uniformly at random. Simulation is halted when there are no more exposed or infected vertices. The quarantine severity *q* is 0.6.

**Figure 9.**
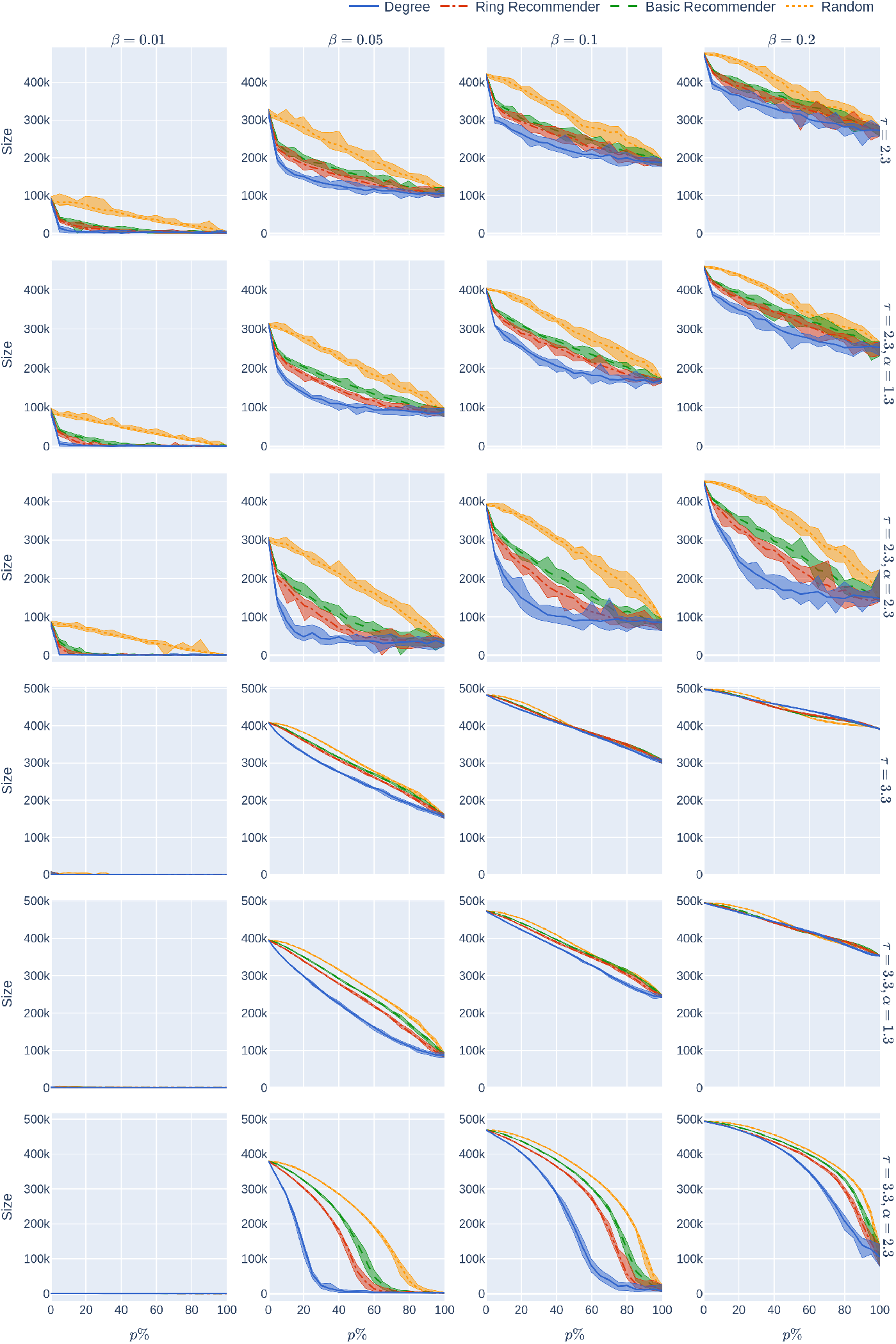
: Epidemic size as a function of the uptake percentage *p*%. The population is modelled using four 500,000-node GIRG networks where *τ* = 2.3 has more super-spreaders and *τ* = 3.3 has fewer super-spreaders, *α* = 1.3 is less geometric and *α* = 2.3 is more geometric, arranged in the four rows. For comparison, there are two rows for 500,000-node configuration models with *τ* = 2.3 and *τ* = 3.3. The *x*-axis shows the uptake percentage *p*% varying from 0 to 100% at step size 5%. Simulations are done for the 21 values of *p*% corresponding to these steps. The *y*-axis shows the corresponding value Size(*p*). Different columns indicate varying infection probabilities *β*. The epidemic size data in Fig 2 corresponds to the *β* = 0.05 column. As in Figure 2, The four uptake scenarios correspond to the four curves on each figure. For each parameter value, the plotted result is the median over 10 runs. The shaded region around the plot covers the results of all 10 simulations. The epidemic is started by infecting 100 individuals, chosen uniformly at random. Simulation is halted when there are no more exposed or infected vertices. The quarantine severity *q* is 0.6.

**Figure 10.**
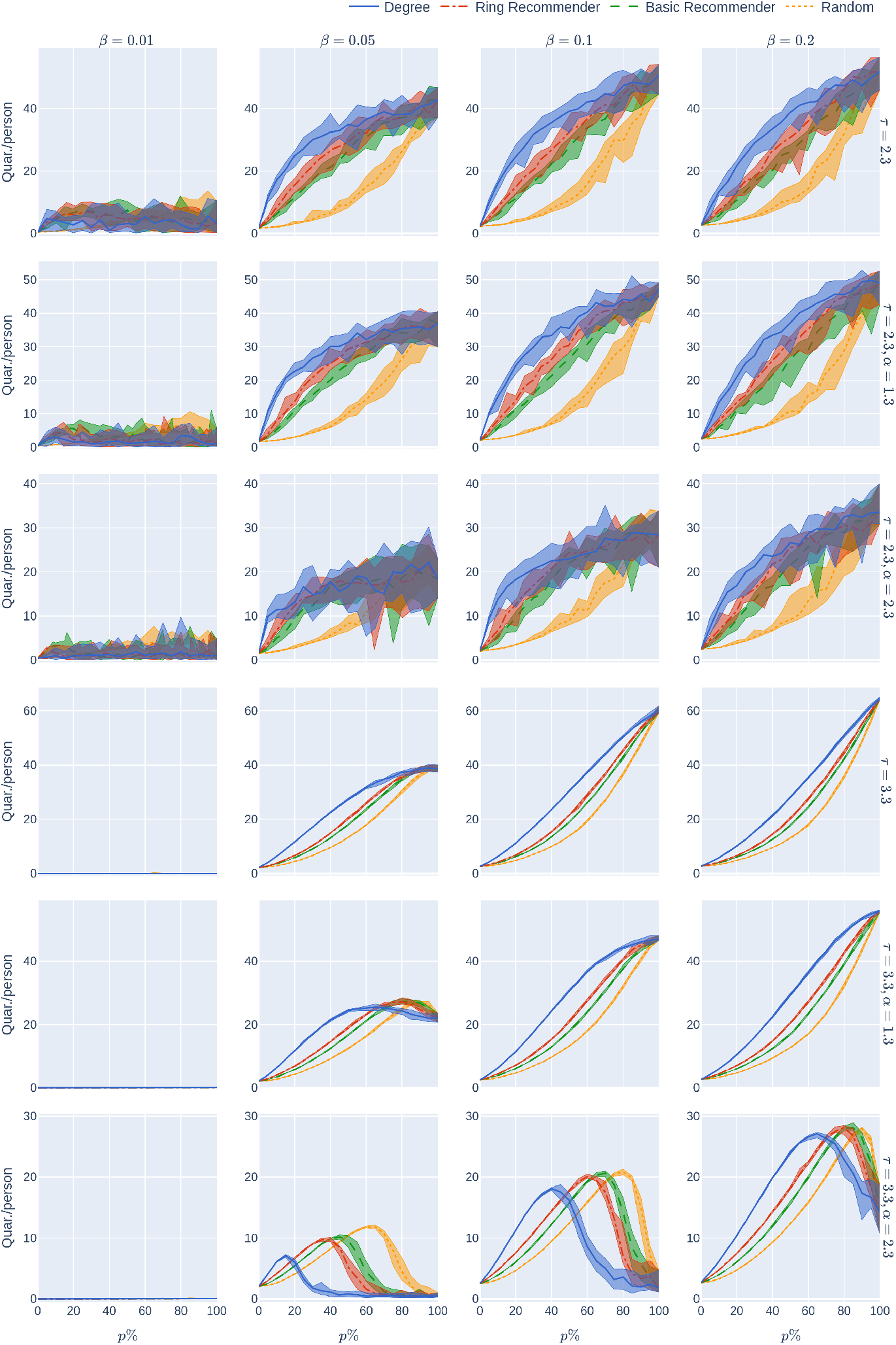
: The average number of days in quarantine per person as a function of the uptake percentage *p*%. The population is modelled using four 500,000-node GIRG networks where *τ* = 2.3 has more super-spreaders and *τ* = 3.3 has fewer super-spreaders, *α* = 1.3 is less geometric and *α* = 2.3 is more geometric, arranged in the four rows. For comparison, there are two rows for 500,000-node configuration models with *τ* = 2.3 and *τ* = 3.3. The *x*-axis shows the uptake percentage *p*% varying from 0 to 100% at step size 5%. Simulations are done for the 21 values of *p*% corresponding to these steps. The *y*-axis shows the corresponding value Quar(*p*). Different columns indicate varying infection probabilities *β*. The Quar data in Fig 2 corresponds to the *β* = 0.05 column. As in Figure 2, The four uptake scenarios correspond to the four curves on each figure. For each parameter value, the plotted result is the median over 10 runs. The shaded region around the plot covers the results of all 10 simulations. The epidemic is started by infecting 100 individuals, chosen uniformly at random. Simulation is halted when there are no more expo2s7ed or infected vertices. The quarantine severity *q* is 0.6.

### 1.4.1 Reducing epidemic size & hospital load by stricter quarantining

Increasing the quarantine-strictness *q* (i.e., requiring individuals to quarantine even with mild symptoms) reduces the number of individuals who are able to spread the virus, and hence reduces the epidemic size and maximum hospital load of the epidemic. This is a very natural observation, valid across all parameter settings and underlying contact networks. The top row of Fig 5 shows how the size of the epidemic decreases, for various CTA-uptake percentages *p*%, as the quarantine-strictness *q* increases. We observe that for *q*_1_ *> q*_2_, for any fixed *p* and uptake scenario □ = rand, basic, ring, deg,

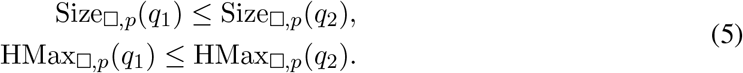

However, the rate of decrease varies across uptake scenarios. Figures 16 and 17 show that in the case with fewer super-spreaders (*τ* = 3.3) typically

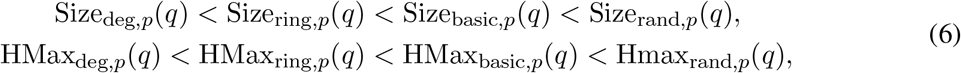

by plotting the various uptake scenarios on the same diagram. To some extent, Equation (6) also holds with more super-spreaders (*τ* = 2.3) but the curves for the recommendation scenarios are close, or overlapping, in these cases (See Figures 18 and 19).

Going back to Figure 5, we find that the Size and HMax curves are steepest under degree-targetted uptake, ring recommendation is second best, while basic recommendation still performs better (declines more steeply) than random uptake. This is similar to what we have already observed in Section 1.3.

Figure 11 is qualitatively similar to Figure 5 and considers an underlying network that is less geometric. For network with more super-spreaders (Figs 12 and 13), these effects are even more exaggerated. It is interesting to observe that the curves of Size_□_(*p*), HMax_□_(*p*) for □ = basic, ring, deg drop so significantly compared to Size_rand_(0), HMqx_rand_(0), respectively, that it appears to be a discontinuity at 0. While we believe these are due to finite size effects, the observation that recommending helps significantly already at very low uptake rates remains valid.

**Figure 11.**
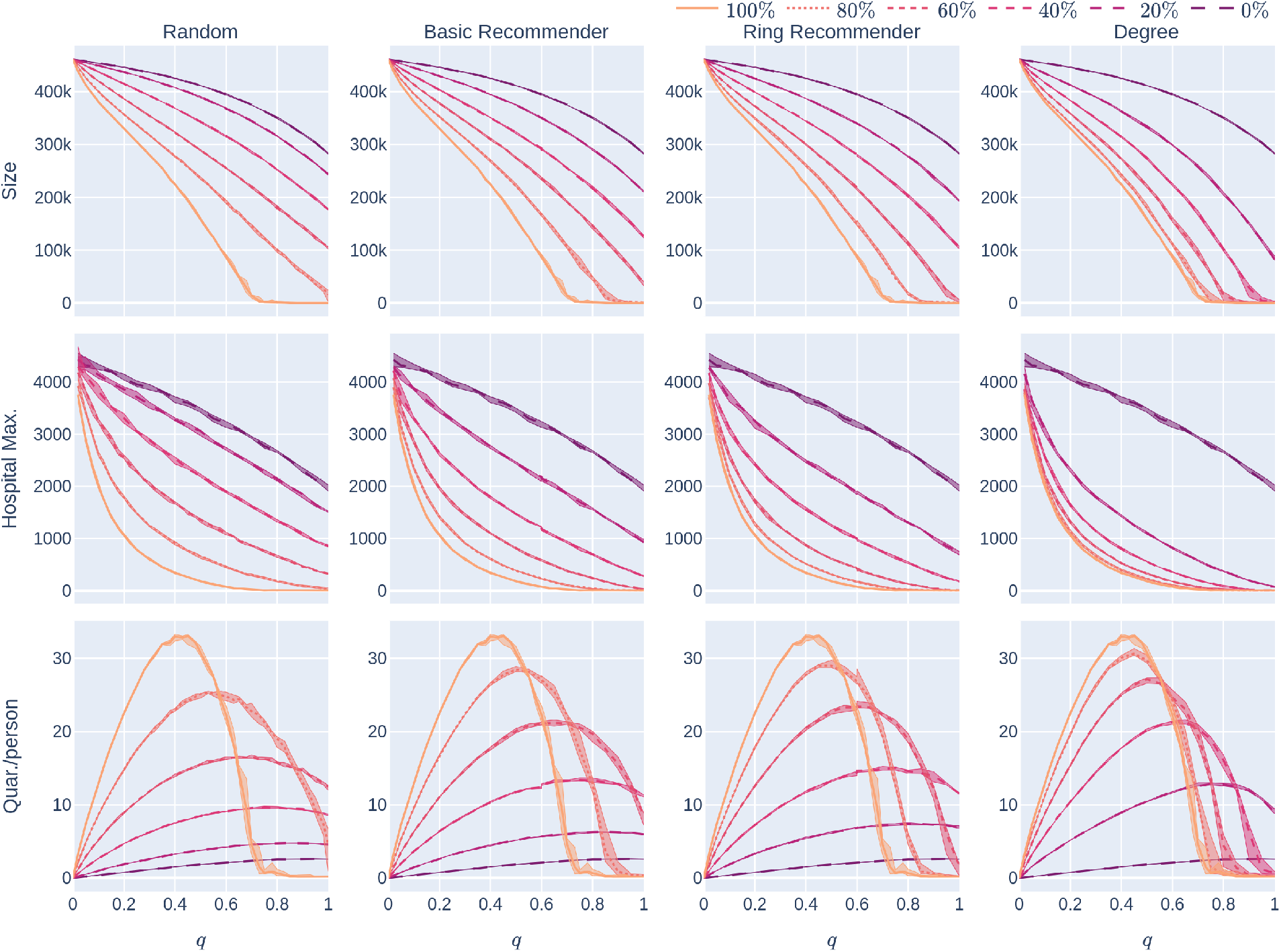
: Experiment 2 for a less geometric network: Influence of the quarantine strictness *q* for various uptake percentages *p*% and the four uptake scenarios: the KPIs are plotted against *q*, the quarantine strictness. We work here with a fixed underlying network, a 500,000 node GIRG with *τ* = 3.3 and *α* = 1.3. Each column represents a given uptake scenario (random, basic-recommender and ring-recommender, and degree-targetted). The *x*-axis shows the quarantine strictness *q* varying from 0 to 1 at step size 0.05. Simulations are done for the 21 values of *q* corresponding to these steps. There is an additional simulation point at 0.02 (in order to avoid division by 0 in the computation for HMax and still have a point close to 0). The *y*-axis shows the corresponding value of the KPI. The curves on each figure correspond to the different uptake percentages, as labelled. For each parameter value, the plotted result is the median over 5 runs. The shaded region around the plot covers the results of all 5 simulations. The epidemic is started by infecting 100 individuals, chosen uniformly at random. Simulation is halted when there are no more exposed or infected vertices. The infection rate is *β* = 0.05.

**Figure 12.**
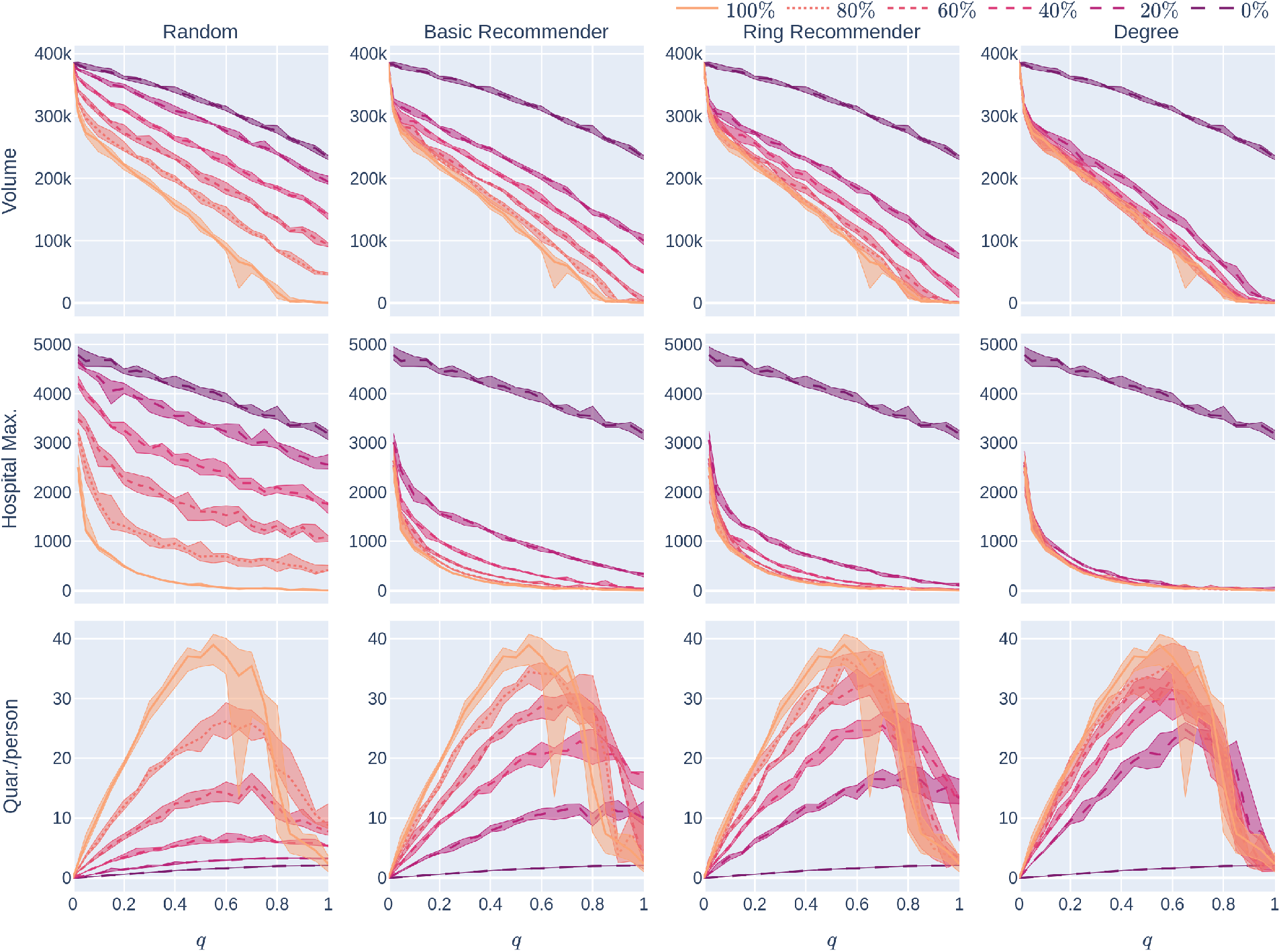
: Experiment 2 on a less geometric network with many super-spreaders: Influence of the quarantine strictness *q* for various uptake percentages *p*% and the four uptake scenarios: the KPIs are plotted against *q*, the quarantine strictness. We work here with a fixed underlying network, a 500,000 node GIRG with *τ* = 2.3 and *α* = 1.3 In this case, the parameter *α* does not have a significant effect since long-range edges are present because of the super-spreaders: the figures are very similar to those in Figure 13. Each column represents a given uptake scenario (random, basic-recommender and ring-recommender, and degree-targetted). The *x*-axis shows the quarantine strictness *q* varying from 0 to 1 at step size 0.05. Simulations are done for the 21 values of *q* corresponding to these steps. There is an additional simulation point at 0.02 (in order to avoid division by 0 in the computation for HMax and still have a point close to 0). The *y*-axis shows the corresponding value of the KPI. The curves on each figure correspond to the different uptake percentages, as labelled. For each parameter value, the plotted result is the median over 5 runs. The shaded region around the plot covers the results of all 5 simulations. The epidemic is started by infecting 100 individuals, chosen uniformly at random. Simulation is halted when there are no more exposed or infected vertices. The infection rate is *β* = 0.05.

**Figure 13.**
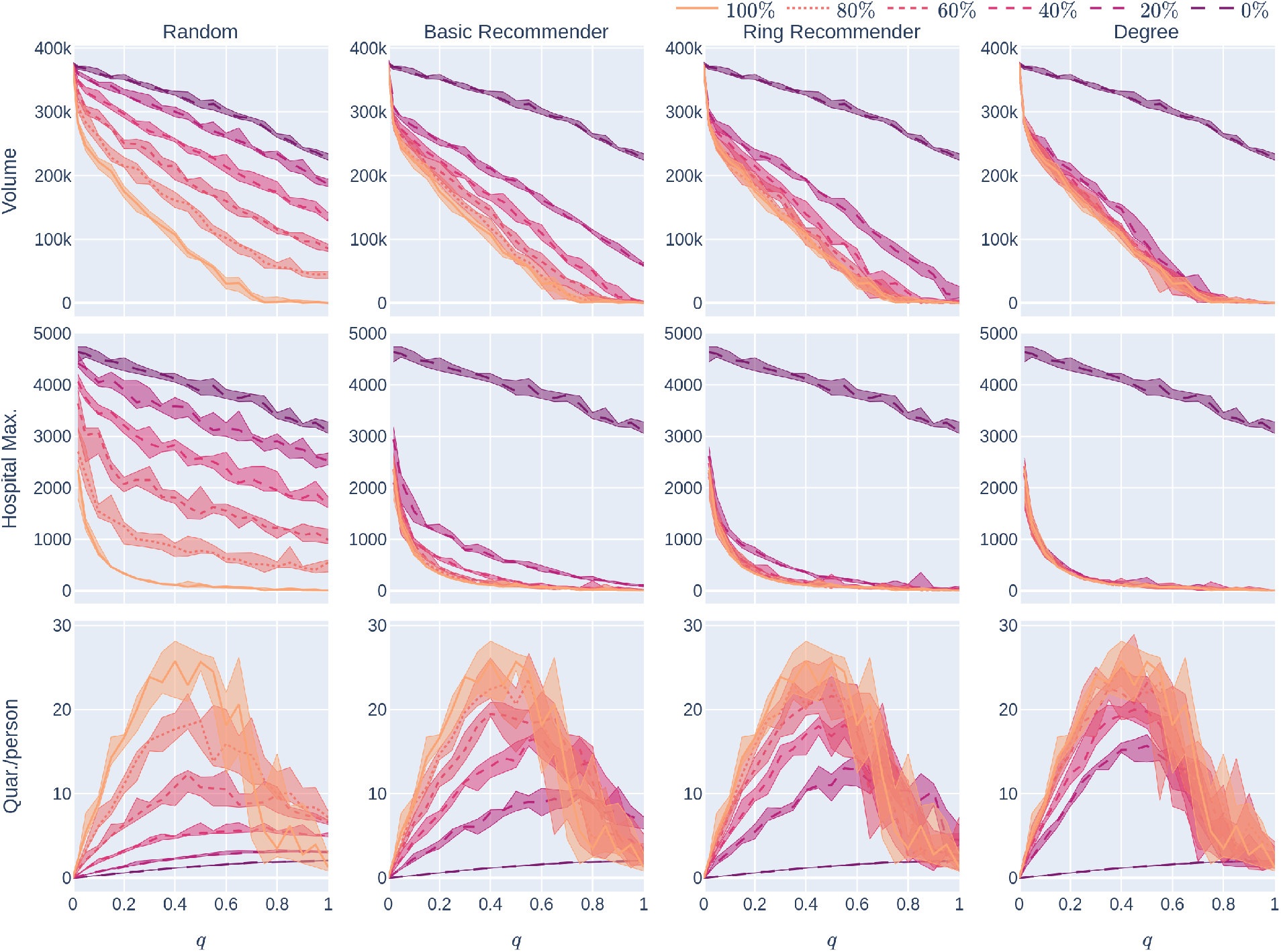
: Experiment 2 on a more geometric network with many super-spreaders: Influence of the quarantine strictness *q* for various uptake percentages *p*% and the four uptake scenarios: the KPIs are plotted against *q*, the quarantine strictness. We work here with a fixed underlying network, a 500,000 node GIRG with *τ* = 2.3 and *α* = 2.3 Observe the similarity with Fig 12. Each column represents a given uptake scenario (random, basic-recommender and ring-recommender, and degree-targetted). Fig 11 below is qualitatively similar and considers an underlying network that is less geometric. The *x*-axis shows the quarantine strictness *q* varying from 0 to 1 at step size 0.05. Simulations are done for the 21 values of *q* corresponding to these steps. There is an additional simulation point at 0.02 (in order to avoid division by 0 in the computation for HMax and still have a point close to 0). The *y*-axis shows the corresponding value of the KPI. The curves on each figure correspond to the different uptake percentages, as labelled. For each parameter value, the plotted result is the median over 5 runs. The shaded region around the plot covers the results of all 5 simulations. The epidemic is started by infecting 100 individuals, chosen uniformly at random. Simulation is halted when there are no more exposed or infected vertices. The infection rate is *β* = 0.05.

### 1.4.2 Improving CTA efficacy by slightly increasing quarantine strength

We next study the shape of the curves Size_□,*p*_(*q*) and HMax_□,*p*_(*q*) for fixed *p* as *q* varies and □ = rand, basic, ring, deg. Intuitively, we examine what happens when we slightly increase the quarantine strength *q*, especially in the mild-quarantine scenario where *q <* 0.5. We first note (From Figure 5) that without a CTA present (*p* = 0%), HMax decreases roughly linearly as a function of *q*. Whenever a CTA is present, (*p*% *>* 0%) HMax becomes a *steeply decreasing convex* function of *q* for low *q*.

The take-home message is: a somewhat stricter quarantine rule can already lower the maximum hospital load and thus “*flatten the curve*” very well in the presence of a CTA. A similar observation is valid for Size_*p*_(*q*): the curve is concave for *p*% = 0%, while it becomes linear or convex when *p* increases.

When the value of *q* is small (so the quarantine measures are not strict), Fig 5 also shows that another way to flatten the curve is to increase the uptake percentage: the rate at which Size and HMax decrease is much higher for large CTA-uptake percentages: our numerical simulations indicate that (for the values of *p* that we have tested) when *p*_1_ *> p*_2_ and *dq* is a small positive number,

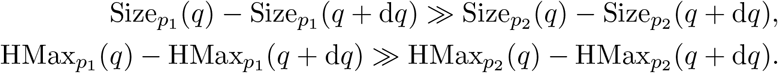

Thus a high(er) CTA uptake rate makes it possible to impose less strict quarantine measures and yet keep the epidemic size and hospital load under control.

We mention that these findings are robust across underlying networks: while Fig 5 shows the KPIs on a fairly geometric network with not too many and not too large super-spreaders, Fig 11 shows the same KPIs for a different class of networks; where many long-range edges are present. The two scenarios are qualitatively similar. For network with more super-spreaders (Figs 12 and 13), these effects are even more exaggerated. It is interesting to observe that the curves of Size_□,*p*_(*q*), HMax_□,*p*_(*q*) for □ = basic, ring, deg drop so significantly compared to Size_□,0_(0), HMAx_□,0_(0), respectively, that it appears to be a discontinuity at 0 in *q*: seemingly, e.g. for *p* = 20%

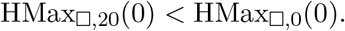

While we believe these are due to finite size effects, the observation that recommending helps significantly already at very low uptake rates remains valid.

### 1.4.3 Reducing quarantining by quarantining

Although it is valuable to increase quarantine-strictness in order to reduce Size and HMax, one may wonder whether stricter quarantine measures require more social sacrifice. Our experiments demonstrate that the picture is not this simple: Row 3 of Fig 5 shows that, for every positive CTA-uptake percentage *p*% and every uptake scenario □ = rand, basic, ring, deg, the curve Quar_□,*p*_(*q*) is roughly unimodal. Thus, once *q* is sufficiently large (larger than some value 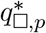 depending on the uptake scenario □ and the uptake percentage *p*%), increasing *q* actually decreases the average number of days that people have to quarantine. One reason for this might be that the quarantining decreases the spread of the epidemic (so other individuals don’t get infected and cause further quarantining).

This observation is most useful if the CTA-uptake percentage *p*% is high enough that 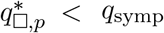, so that quarantine-strictness quantities 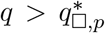 are easy to implement in reality. Note that 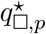 also depends on the infectiousness of the disease. Stricter social distancing, corresponding to lower reproduction number, leads to lower value of 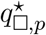. Thus, when social-distancing measures are in place, it will be more possible to reduce the average time that people quarantine by setting 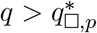.

### 1.4.4 Geometry helps

Fig 14 shows the effect of geometry on the epidemic size and maximum hospital load as a function of the quarantine strictness function *q*, for two levels of infectiousness (high and low). Observe that networks with more geometry (*α* = 2.3) have lower epidemic size and lower maximum hospital load than networks with many long-range edges and hence less geometry (*α* = 1.3), in this respect, the conclusion is similar to that in Experiment 1, see Section 1.3.3.

**Figure 14.**
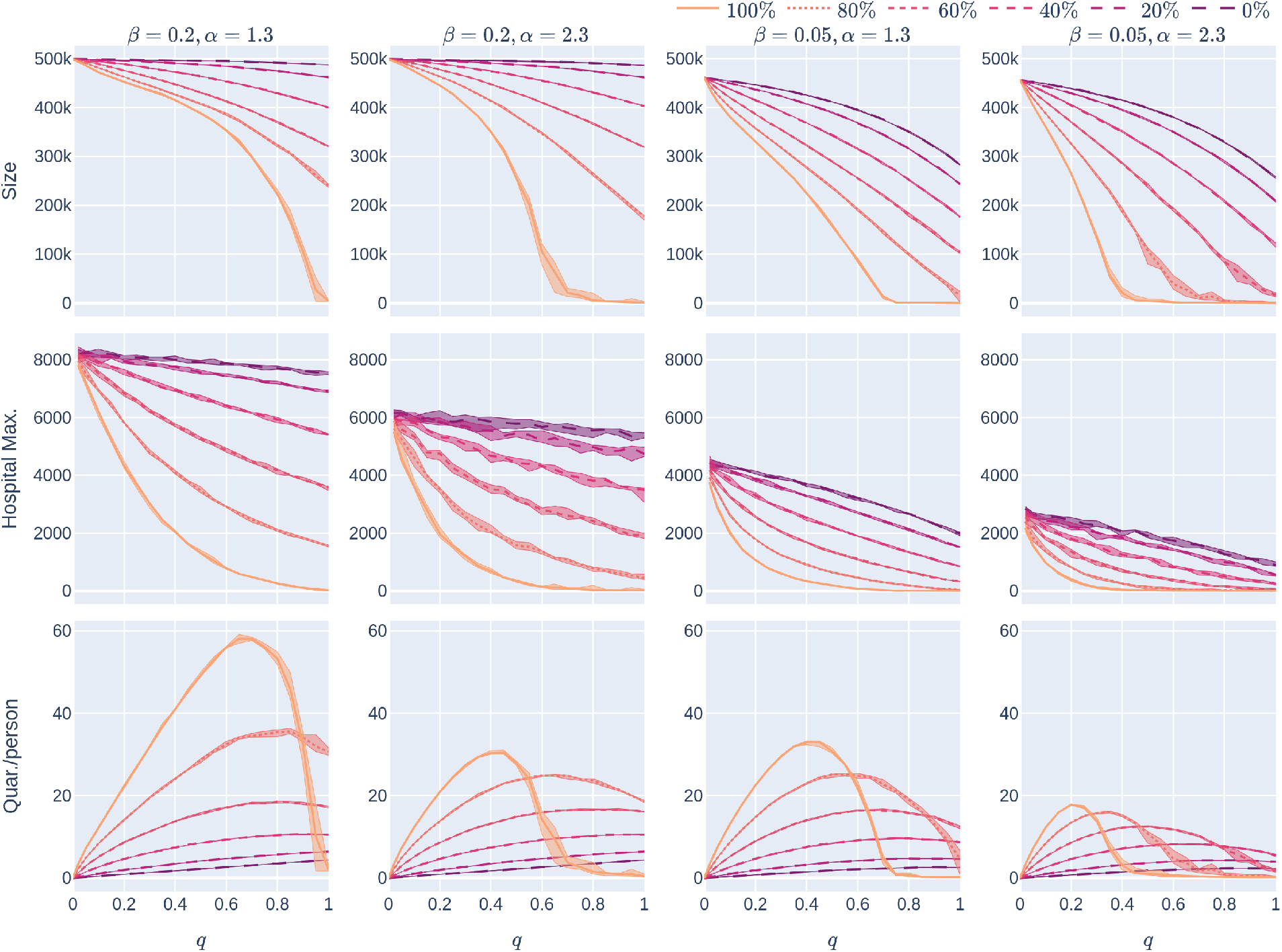
: The effect of geometry and level of infectiousness on Experiment 2: The different curves correspond to different CTA uptake-rates (*p*%), all in the random-uptake scenario in a 500,000-node GIRG with few super-spreaders (*τ* = 3.3). The four columns represent different scenarios – the first column has *β* = 0.2 (a high infection rate, perhaps caused by less social distancing) and *α* = 1.3 (a less geometric network). The second column also has *β* = 0.2, but with more geometry (*α* = 2.3). The final two columns represent scenarios with a lower underlying infection rate (*β* = 0.05), with varying levels of geometry. The three rows correspond to the three KPIs. The *x*-axis shows the quarantine strictness *q* varying from 0 to 1 at step size 0.05. Simulations are done for the 21 values of *q* corresponding to these steps. There is an additional simulation point at 0.02 (in order to avoid division by 0 in the computation for HMax and still have a point close to 0). For each parameter value, the plotted result is the median over 5 runs. The shaded region around the plot covers the results of all 5 simulations. The epidemic is started by infecting 100 individuals, chosen uniformly at random. Simulation is halted when there are no more exposed or infected vertices.

Fig 15 shows that in case there are many super-spreaders in the network (*τ* = 2.3), these form a well-connected core so that the value of *α* becomes less significant: in this case geometry is not well described by this parameter. See more on this in Section 3.1 below.

**Figure 15.**
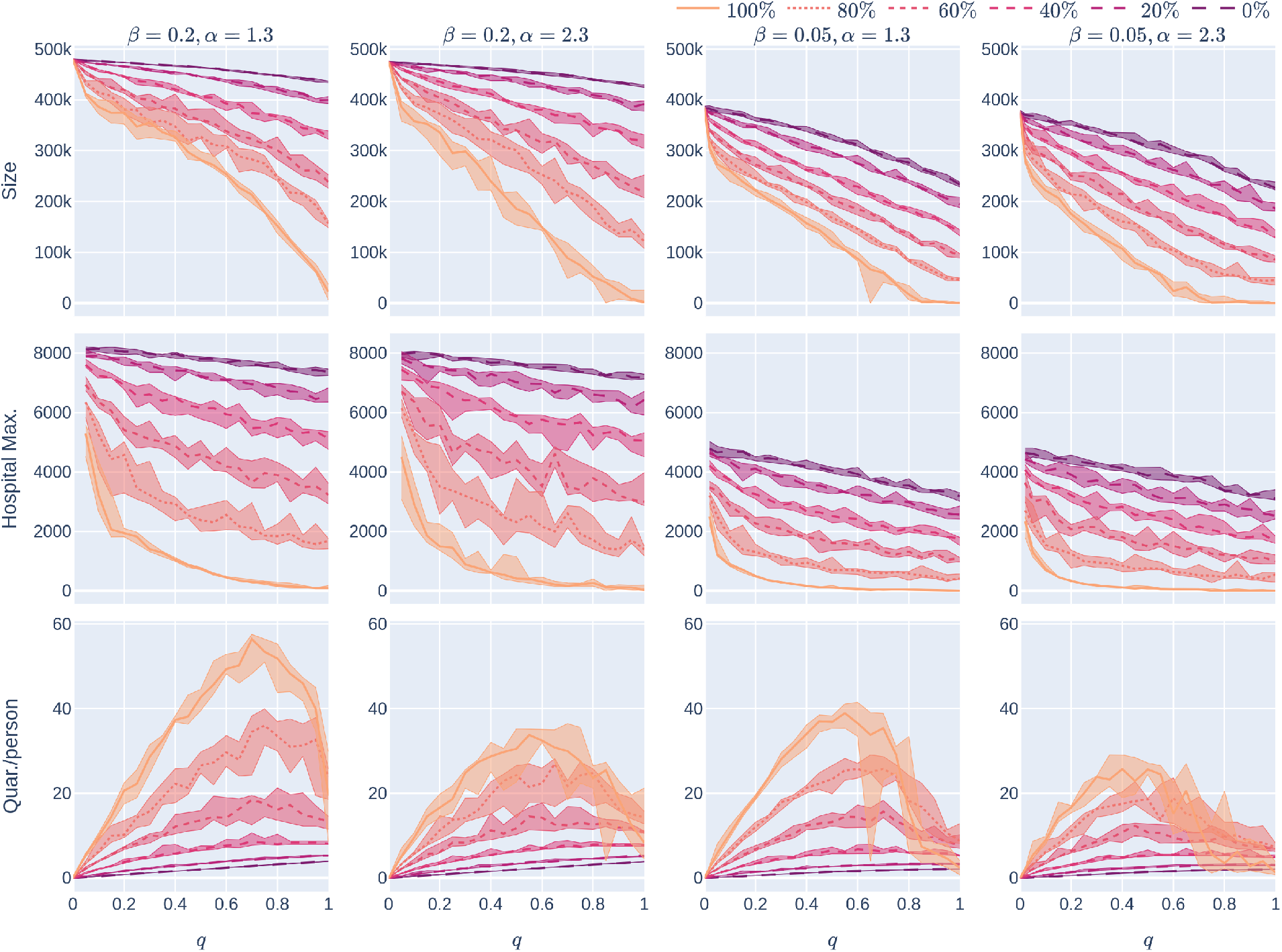
: The effect of geometry and level of infectiousness on Experiment 2: The different curves correspond to different CTA uptake-rates (*p*%), all in the random-uptake scenario in a 500,000-node GIRG with many super-spreaders (*τ* = 2.3). The four columns represent different scenarios – the first column has *β* = 0.2 (a high infection rate, perhaps caused by less social distancing) and *α* = 1.3. The second column also has *β* = 0.2, but with a higher *α*, implying fewer (but not significantly fewer) long-range edges. Observe that the effect of increasing *α* is not very significant, corresponding to the explanation in Section 1.4.4. The final two columns represent scenarios with a lower underlying infection rate (*β* = 0.05), with varying levels of geometry. The three rows correspond to the three KPIs. The *x*-axis shows the quarantine strictness *q* varying from 0 to 1 at step size 0.05. Simulations are done for the 21 values of *q* corresponding to these steps. There is an additional simulation point at 0.22 (in order to avoid division by 0 in the computation for HMax and still have a point close to 0). For each parameter value, the plotted result is the median over 5 runs. The shaded region around the plot covers the results of all 5 simulations. The epidemic is started by infecting 100 individuals, chosen uniformly at random. Simulation is halted when there are no more exposed or infected vertices.

**Figure 16.**
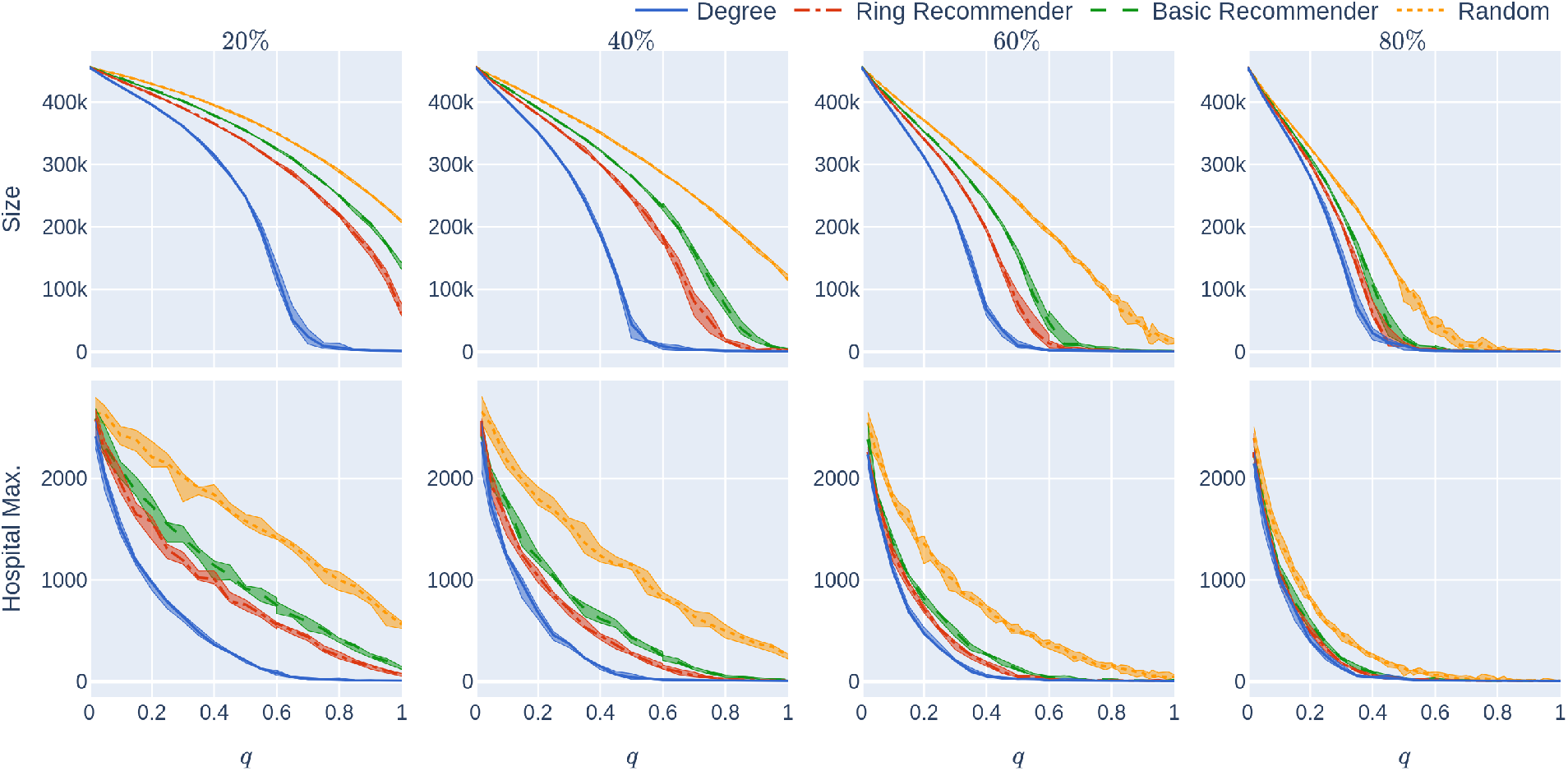
: Experiment 2 on a geometric network with not too many super-spreaders: Influence of the quarantine strictness *q* for various uptake percentages *p*% and the four uptake scenarios at infection rate *β* = 0.05. The underlying network is a 500,000-node GIRG with *τ* = 3.3 (fewer super-spreaders) and *α* = 2.3 (more geometric). Each column represents an uptake percentage. The *x*-axis shows the quarantine strictness *q* varying from 0 to 1 at step size 0.05. Simulations are done for the 21 values of *q* corresponding to these steps. There is an additional simulation point at 0.02 (in order to avoid division by 0 in the computation for HMax and still have a point close to 0). The *y*-axis shows the corresponding value of the KPI. The curves in each figure correspond to the different uptake scenarios, as labelled. For each parameter value, the plotted result is the median over 5 runs. The shaded region around the plot covers the results of all 5 simulations. The epidemic is started by infecting 100 individuals, chosen uniformly at random. Simulation is halted when there are no more exposed or infected vertices.

**Figure 17.**
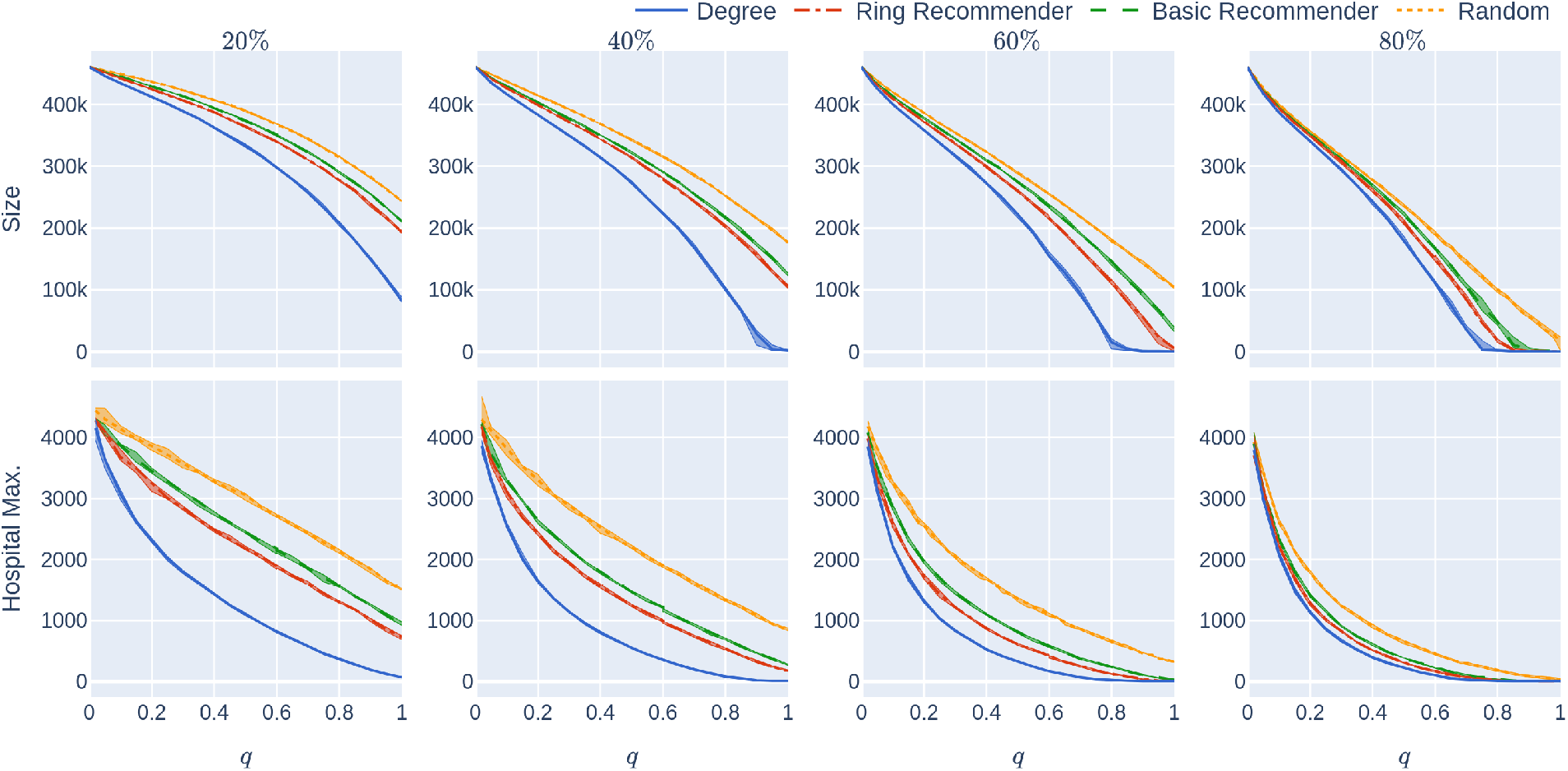
: Experiment 2 on a less geometric network with not too many super-spreaders: Influence of the quarantine strictness *q* for various uptake percentages *p*% and the four uptake scenarios at infection rate *β* = 0.05. The underlying network is a 500,000-node GIRG with *τ* = 3.3 (fewer super-spreaders) and *α* = 1.3 (less geometric). Each column represents an uptake percentage. The *x*-axis shows the quarantine strictness *q* varying from 0 to 1 at step size 0.05. Simulations are done for the 21 values of *q* corresponding to these steps. There is an additional simulation point at 0.02 (in order to avoid division by 0 in the computation for HMax and still have a point close to 0). The *y*-axis shows the corresponding value of the KPI. The curves in each figure correspond to the different uptake scenarios, as labelled. For each parameter value, the plotted result is the median over 5 runs. The shaded region around the plot covers the results of all 5 simulations. The epidemic is started by infecting 100 individuals, chosen uniformly at random. Simulation is halted when there are no more exposed or infected vertices.

**Figure 18.**
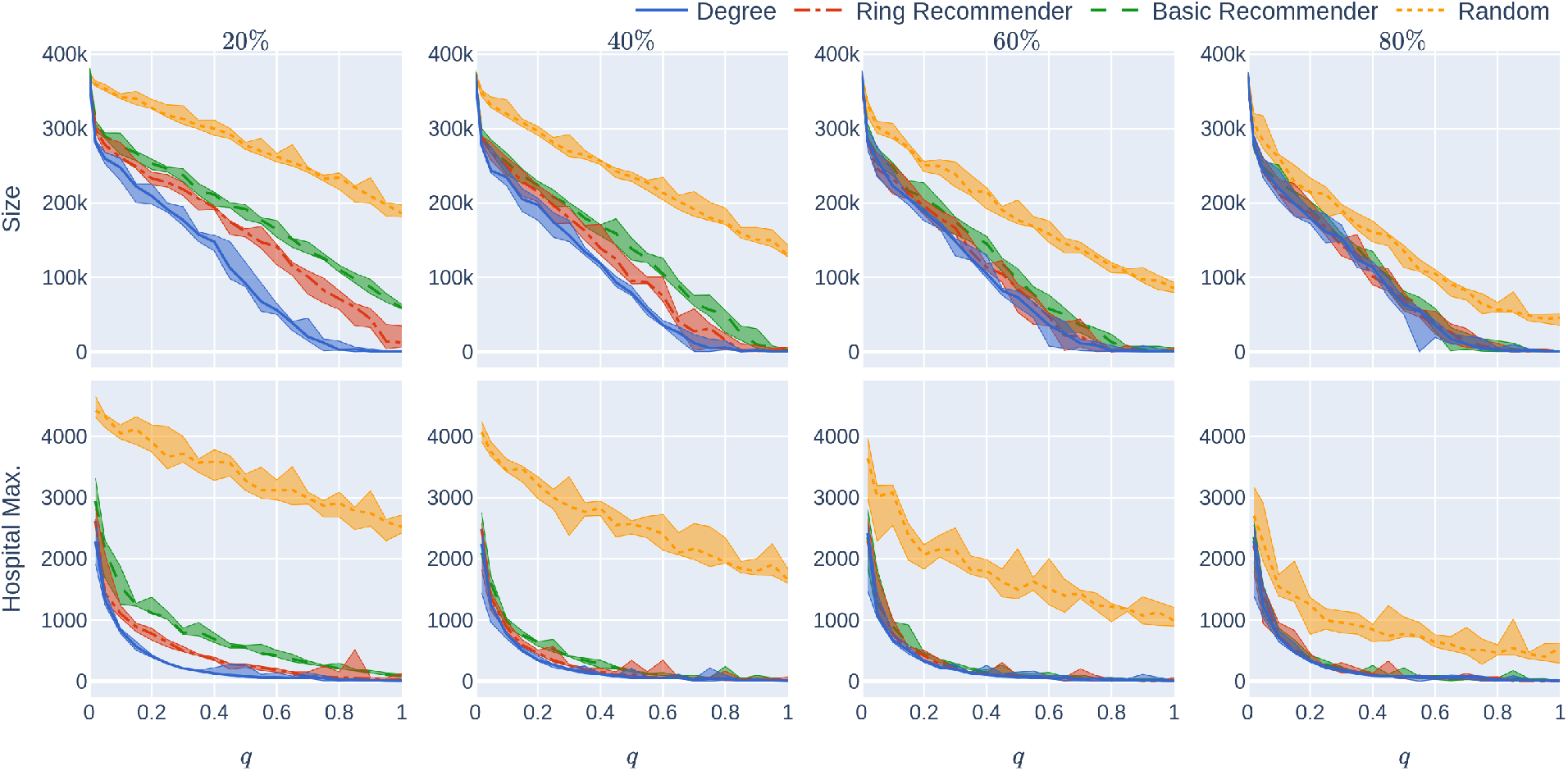
: Experiment 2 on a geometric network with many super-spreaders: Influence of the quarantine strictness *q* for various uptake percentages *p*% and the four uptake scenarios at infection rate *β* = 0.05. The underlying network is a 500,000-node GIRG with *τ* = 2.3 (more super-spreaders) and *α* = 2.3 (more geometric). Each column represents an uptake percentage. The *x*-axis shows the quarantine strictness *q* varying from 0 to 1 at step size 0.05. Simulations are done for the 21 values of *q* corresponding to these steps. There is an additional simulation point at 0.02 (in order to avoid division by 0 in the computation for HMax and still have a point close to 0). The *y*-axis shows the corresponding value of the KPI. The curves in each figure correspond to the different uptake scenarios, as labelled. For each parameter value, the plotted result is the median over 5 runs. The shaded region around the plot covers the results of all 5 simulations. The epidemic is started by infecting 100 individuals, chosen uniformly at random. Simulation is halted when there are no more exposed or infected vertices.

**Figure 19.**
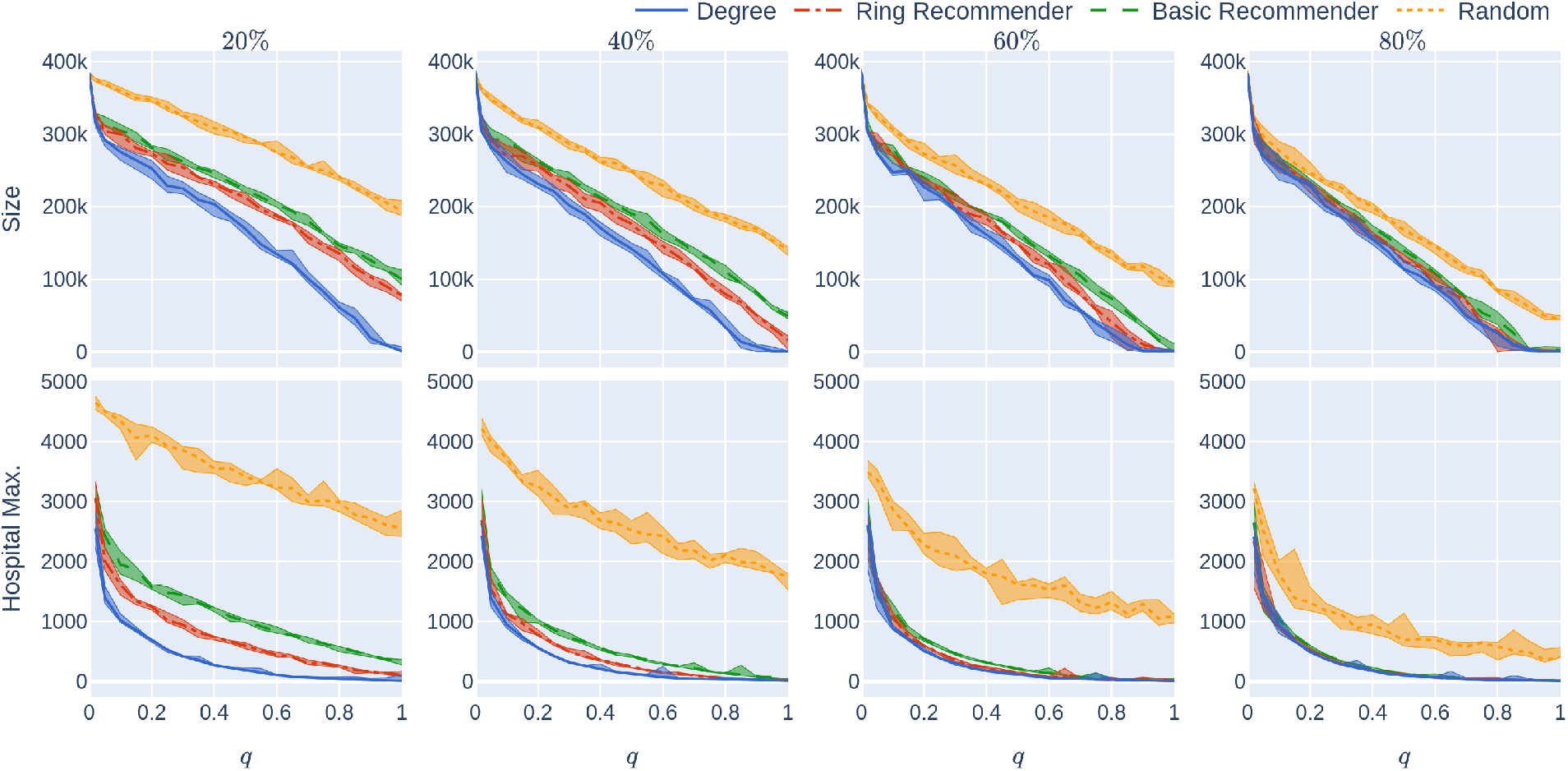
: Experiment 2 on a less geometric network with many super-spreaders: Influence of the quarantine strictness *q* for various uptake percentages *p*% and the four uptake scenarios at infection rate *β* = 0.05. The underlying network is a 500,000-node GIRG with *τ* = 2.3 (more super-spreaders) and *α* = 1.3 (less geometric). Each column represents an uptake percentage. The *x*-axis shows the quarantine strictness *q* varying from 0 to 1 at step size 0.05. Simulations are done for the 21 values of *q* corresponding to these steps. There is an additional simulation point at 0.02 (in order to avoid division by 0 in the computation for HMax and still have a point close to 0). The *y*-axis shows the corresponding value of the KPI. The curves in each figure correspond to the different uptake scenarios, as labelled. For each parameter value, the plotted result is the median over 5 runs. The shaded region around the plot covers the results of all 5 simulations. The epidemic is started by infecting 100 individuals, chosen uniformly at random. Simulation is halted when there are no more exposed or infected vertices.

### 1.4.5 Other parameter values

The experiments in Fig 5 assumed an epidemic model (see Fig 2) with a low rate of infection *β* = 0.05. As we noted earlier, low rates of infection can be achieved by social-distancing measures. Fig 14 repeats the experiment for other values of *β*.

### 1.5 Remark: Social distancing and basic reproduction number

As one would expect, social-distancing measures, which reduce the probability that any particular interaction leads to an infection, increase the effectiveness of CTAs. This is observed in our experiments — for example, monotonicity in *β* can be observed by comparing the columns in Figures 8 and 9.

## 2 The epidemic model

The epidemic model that we use is a refinement of the agent-based version of the discrete-time SEIR model. The spreading process changes at discrete time steps, *t* = {0, 1, 2, …}, each time step corresponds to, say, a day. We fix the network *G* in advance. We think of nodes in the network as individuals, and denote the set of nodes by 𝒱. The neighbours of a node *u* are nodes with a direct connection (also called link, or edge) to *u*. A connection may correspond to a friendship or an acquaintance, or simply a contact event.

As we explained in the context of Experiment 2, the model has a parameter *q* ∈ [0, 1] corresponding to quarantine-strictness. The quantity *q* is the fraction of individuals whose symptoms would be so severe that they would go into home-quarantine if infected. As we already explained, determining the value of *q* is a social/economic decision. The model reflects this decision as follows: For each node, independently, and in advance of the epidemic simulation, it is determined randomly, with probability *q*, whether the individual corresponding to this node will, upon infection, develop symptoms exceeding the socially-determined symptom severity threshold, and then go into home-quarantine. The set of nodes which will do so is called 𝒱_sev_ and the set of nodes which will not is called 𝒱_mild_. During the epidemic, nodes in 𝒱_sev_ that become infected will go into home-quarantine. One may worry about uncooperative individuals who do not self-isolate, despite having sufficiently severe symptoms. We do not need to adjust to model to account for these, since the quarantine-strictness *q* can be viewed as the fraction of individuals that (1) would develop sufficiently-severe symptoms, and (2) would be willing to self-isolate. Note that the actual distribution of the symptom-strength in the population is irrelevant to the problems that we study; the only relevant parameter is the final proportion *q*. The assumption that we do we make, however, is that each individual has an independent symptom-strength variable, making 𝒱_sev_ a random set of nodes.

### Definition 1

(SEISeR model). *Determine the node sets* 𝒱_sev_ *and* 𝒱_mild_. *Nodes in* 𝒱_sev_ *can be in five possible states:* susceptible (S), exposed (E), infectious but not yet showing severe symptoms (I), infectious and showing severe symptoms (Se), or removed (R), *while nodes in* 𝒱 _mild_ *can only be in state* (*S*), (*E*), (*I*) *or* (*R*). *At t* = 0, *a subset of nodes ε*_0_⊂ 𝒱 *is put in state (E), all other nodes are in state* (*S*) *(susceptible). The discrete time dynamics between the states are described as follows (see also Fig 6):*

- **Infecting:** *Each infectious node (in state (I))*, infects *each of its susceptible neighbours within the network with probability β at every time step. Infections to different neighbours happen independently. These neighbour nodes enter state (E), exposed to the virus*.
- **Exposed** → **Infectious (not (yet) severely symptomatic)** *When exposed to the virus, each node becomes infectious (transitions to state* (*I*)*) with probability γ at every time step, independently of other nodes*.
- **Infectious** → **Severely Symptomatic** *When a node in* 𝒱_sev_ *is in state (I), (infectious but does not show severe symptoms yet), it transitions to state (Se) with probability γ*_sev_ = 1*/*2.5 *at each time step*.
- **Infectious** → **Removed** *When a node in* 𝒱_mild_ *is in state (I), infectious but does not show severe symptoms, it heals and transitions to state (R) with probability η*_*mild*_ = 1*/*7 *at each time step*.
- **Severe Symptomatic** → **Removed** *When a node is showing severe symptoms, it* heals *with probability η*_*sev*_ = 1*/*4.5 *at every time step, independently of other nodes. Upon healing, the node enters state R (removed)*.

We emphasise that in our model, severely symptomatic nodes do not infect anymore. We assume that they self-quarantine until being removed. In reality, some infectious individuals may choose not to self-quarantine, but we have already incorporated this into the model Recall that 𝒱 _sev_ contains only nodes that would develop sufficiently severe symptoms, and would go into home-quarantine.

To keep the parameter space tractable, we set *γ* = 1*/*3, *η*_mild_ = 1*/*7, *γ*_sev_ = 1*/*2.5, and *η*_sev_ = 1*/*4.5. Thus, in the model, after being exposed to the virus, it takes on average 3 days to become infectious and a further 7 days to heal completely for each individual. Nodes in 𝒱_sev_ infect for an average of 2.5 days before showing symptoms and moving to isolation (that lasts until their perfect healing) while nodes in 𝒱_mild_ are infectious for an average of 7 days before removal. These values are based on empirical findings e.g. in [30, 31, 1, 32], where we emphasise that our results are robust in these parameters. Experiment 2 varies the value of *q*, which is the expectation of |𝒱_sev_| */* |𝒱|.

Next, we describe our model for the CTA. First we give the details of the four CTA-uptake scenarios from Section 1.3 then we define the necessary modifications to the dynamics of the SEISeR model above to incorporate the presence of a CTA.

### Definition 2

(Random, basic-recommender, ring-recommender, and degree-targetted CTA uptake). *Before the epidemic starts, we determine the sets* 𝒱_*u*_, 𝒱_*nu*_ *of users and non-users of the application. We denote by p* = 100 | 𝒱_*u*_ | */* | 𝒱 | *the empirical uptake percentage. We say that the user uptake is*

- **Random**: *When the nodes that use that application are chosen uniformly at random*.
- **Degree-targetted**: *When the p ·* |𝒱| */*100 *nodes with the highest degree (number of connections) are targetted to use the CTA*.
- **Basic-Recommender:** *When an initial set of nodes is chosen uniformly at random. Each node in this initial set uses the CTA, and also recommends it to one of its neighbour chosen uniformly at random, who will also use the CTA. The resulting number of users is p* · |𝒱|*/*100
- **Ring-Recommender:** *When an initial set of nodes is chosen uniformly at random. Each node in this initial set uses the CTA, and also recommends it to all of its neighbour, half of whom (chosen uniformly at random) will also use the CTA. Again, the resulting number of users if p* · |𝒱|*/*100.

We observe that degree-targetted uptake is an unrealistic scenario to consider, since it requires a complete knowledge of the contact network. However, studying the performance of this hypothetical scenario gives a baseline comparison for determining the potential performance of a given uptake percentage. We also emphasise that in the basic-recommendation and ring-recommendation uptake scenarios, the performance of the final uptake percentage *p*% = 100 | 𝒱_*u*_ | */* | 𝒱 | % is compared to the performance of other uptake scenarios with the same *p*%. In other words, the basis for comparison is not the proportion of the initially chosen set. Instead, it is the final uptake percentage. See Fig 1 for the relation between the initial and final uptake percentage for the four networks.

Next, we describe the necessary modifications to the SEISeR model in the presence of a CTA. Informally, a CTA user, upon either being tested positive or simply showing sufficient symptoms, will notify its contacts to stay in quarantine for a duration of *T* days (*T* = 14 in most countries), regardless of whether these contacts themselves show symptoms.

### Definition 3

(SEISeR with a contact-tracing application (CTA)). *In addition to the original states (S), (E), (I), (Se) and (R) of the SEISeR model, there are five new states (NS), (NE), (NI), (NSe) and (NR), corresponding to “notified” versions of the original states — we refer to these as “N -states” and we refer to the original states as “O-states”. Only CTA-users (nodes in* 𝒱_u_*) will ever enter the N -states. They enter the N -states when they receive a notification, via the CTA. They stay in the N -states until T* = 14 *days have elapsed since the last notification received. While in N -states, they self-quarantine, so cannot spread infection*.

*The transitions between the N -states are exactly the same as the transitions between the original versions of these states, except that there is no transition from (NS) to (NE), because the notified nodes in state (NS) will be quarantining, so will not become exposed. See the black arrows denoting transitions in Fig 7*.

*All nodes start in O-states, exactly as in the SEISeR model. After each time step t, nodes move between the O-states and the N -states as follows:*

- **Notification:** *If a node in* 𝒱_*u*_ *starts to show sufficient symptoms at time t (i*.*e*., *either it transitions from (I) to (Se) at time t or it transitions from (NI) to (NSe) at time t) then it sends a notification to all of its network-neighbours that are in* 𝒱_*u*_.
- **Moving to** *N* **-states:** *If a node in an O-state receives a notification, it moves to its corresponding N -state (following the corresponding red arrow in Fig 7)*.
- **Moving to** *O***-states:** *If a node is in an N -state and it has not received a notification for T* = 14 *time steps, then it moves to its corresponding O-state (following the corresponding blue arrow in Fig 7)*.

### Justification of modelling choices and robustness

We make a few comments to justify our modelling choices. Nodes in the *N* -states and in state (Se) are assumed to be self-quarantining, so they cannot spread infection. These are the blue nodes in Fig 7). Only nodes in state (I) (coloured red) can spread infection.

#### Sending Notifications

The reason that nodes entering state (NSe) send notifications (even though they are already self-quarantining) is that they may have spread infection before entering the *N* -states (and starting to quarantine): their neighbours may be in exposed or infectious states when receiving the notification. In fact, while this model does not include testing, such a scenario also happens in real life when an individual, already part of a contact tracing chain, receives a positive test. In this case, the rest of the contacts of this individual need to be notified as well.

#### Removed nodes in quarantine

The underlying SEISeR model assumes that individuals can only be infected once (so “removed” nodes do not become susceptible again). Despite this, in light of possible re-infection, many countries (e.g. the Netherlands, where two of the authors are located) do ask individuals who are notified to self-isolate, even if they have already have Covid-19. While transitioning from (R) to (NR) has no effect on the course of the epidemic, it does have an effect on the total population’s effective workforce (which we also study). Indeed, the self-isolating blue nodes represent a social cost, since all of these nodes are self-isolating, and cannot go to work. A node who is say, exposed when it receives a notification via a CTA may go through all the phases (NE), (NI), (NSe), (NR) before the *T* days are over. In this case it cannot become simply removed and go back to work immediately, but has to wait out the *T* days and then transition to state (R). For the same reason we make a distinction between nodes in (Se) and (NSe): nodes in (Se) may leave home isolation immediately after healing, while nodes in (NSe) have to wait for the necessary total *T* days even after healing to be able to leave isolation.

#### Multiple notifications extend the quarantine time

We make a further comment on why we choose to extend the quarantine by *T* days after an additional notification is sent to a node already in an *N* -state. The reason for this is again based on real life experience: since not all contacts and hence notifications lead to infection, a second notification should not be ignored. Whether individuals are actually willing to comply with such rules belongs to behavioural science and is out of the scope of the current study. We assume an idealised scenario where each node in 𝒱_*u*_ complies with the rules. Varying the size of *𝒱*_*u*_ captures the effect of less compliance as well.

#### Robustness of choices

To conclude the justification, we emphasise that we conduct a *qualitative and comparative* study. The model is sufficiently robust that the conclusions of our study also apply under small changes to the model. For example, if an uptake scenario performs better than another with regard to this particular model, it will also perform better in a model with slight changes, e.g. a model that does not extend the time of home isolation when a second notification is received, or a model that does not require already-removed nodes to go into quarantine.

We note that since each infectious node infects each of its susceptible neighbours with probability *β* at each time step, this model is a reactive process in the sense of [33]. Next we describe the underlying contact network of the agent-based epidemic.

## 3 Agent-based population model: spatial scale free network models

To be able to carry out our study, we represent the underlying contact network of individuals (nodes, or agents) as a network. Based on the universality results in [21], our primary choice for the underlying contact network is a mixture of pure geometric and purely random network models, called Geometric Inhomogeneous Random Graphs, which possess geometric features and can match the statistical properties of real human contact networks, such as degree distributions and clustering, as well as longrange connections. For baseline comparison, we also study the case when the underlying network is a “mean-field network”, ignoring the spatial component. For this scenario we use the *configuration model*, which can mimic the local statistical properties of real human contact networks, such as degree distributions.

### 3.1 Geometric Inhomogeneous Random Graphs

We model populations embedded in geometric space using Geometric Inhomogeneous Random Graphs (GIRGs). Recently, several spatial random graph models were developed to mimic properties of real networks features: hyperbolic random graphs [34, 35, 36], scale-free percolation [37], and GIRGs [38, 4, 39]. The qualitative behaviour of these models is the same, in fact, they can be unified into a general model containing all three models as special cases. For sake of simplicity we decided to work with GIRGs.

The advantage of these spatial models are that they can mimic several aspects of real contact networks: individuals are embedded in space, just like in real life, allowing for local community structures to be present in the contact network. This results in strong clustering [39, 40]. Beyond mimicking the degree distribution of real networks (e.g. high variability) GIRGs also incorporate connections (edges) bridging spatial distance on all scales, short as well as long-range edges. The model is very flexible, and for properly chosen parameter settings it is *scale-free* in two respects: both in spatial distance that edges cover, and in node-degree variability [39].

Contactor activity networks of humans have been found to show similar behaviour, including heavy-tailed degree distributions, strong clustering, and community structures [41, 42, 43, 44, 45], as well as heavy-tailed distance-distribution for edges, see references in [46]. For epidemic spread specifically, lighter tailed degree distributions such as negative binomial have been observed e.g. in [1, 31]. GIRGs also allow for the incorporation of such distributions, by setting light-tailed fitness distributions.

The following definition from [21] is general, and in its last sentence we specify it for a GIRG. The underlying space can be the earth’s surface, or ℝ^2^, the two-dimensional Euclidean space. We denote by *x* ∧ *y* the minimum of two numbers *x, y*.

#### Definition 4

(Geometric Inhomogeneous Random Graph (GIRG)). *Fix N* ≥ 1 *the number of nodes. Assign to each node u* ∈ {1, 2, …, *n*} *a* fitness *w*_*u*_ *>* 0, *and a* spatial location Φ(*u*). *Fix α >* 0. *For any pair of nodes u, v with fixed w*_*u*_, *w*_*v*_, Φ (*u*), Φ (*v*), *connect them by an edge with probability*

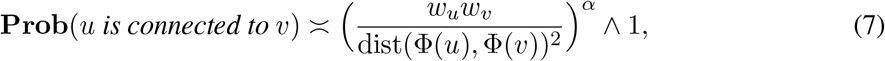

*where* dist *is a distance function, i*.*e*., *it measures the distance between the locations* Φ(*u*) *and* Φ(*v*). *When* Φ(*u*) *is chosen uniformly in a box of volume N, we obtain a GIRG*.

Mathematically, the distance function dist in Definition 4 is a metric on the underlying space, (e.g., a metric such as ‖ · ‖_2_). In the definition, the formula is given for a two-dimensional model. In higher dimensions, the exponent 2*α* in the denominator would change to dim · *α*, with dim being the dimension of the model.

GIRGs have a natural interpretation: the fitnesses express the ability of nodes to have many connections, Φ embeds them in space, and *α* is the *long-range* parameter: the smaller *α* is, the more the model favours longer connections. The parameter space of GIRG is rich enough to model many desired features observed in real networks:

a. extreme variability of the number of neighbours (degrees),
b. connections present on all length-scales,
c. small and ultra-small distances,
d. strong clustering,
e. local communities.

In real-life networks, an extreme variability of node degree is often observed, see [41, 47, 44]. Extreme node degree variability results in the presence of a few individuals with extreme influence on spreading processes, the *hubs or super-spreaders* [44]. Mathematically, this extreme degree variability can be expressed using the empirical distribution of node degrees, that follows a *power-law*:

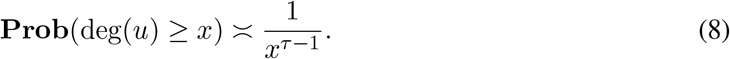

for some *τ >* 2. Setting a power-law fitness distribution for *w*_*u*_ in a GIRG yields that the degrees satisfy (8). As we shall see, the parameter *α* in (7) determines the presence of long-range connections: as *α* gets smaller, it is more likely that there are long-range connections. Thus, the two parameters of the model are *τ* and *α*.

The parameters *τ* and *α* determine the average graph distance between nodes in a GIRG [48, 38, 37, 49]. Letting *d*_*G*_(*u, v*) denote the graph distance between *u* and *v* (the number of connections on the shortest path between nodes *u* and *v*), the average distance is given by

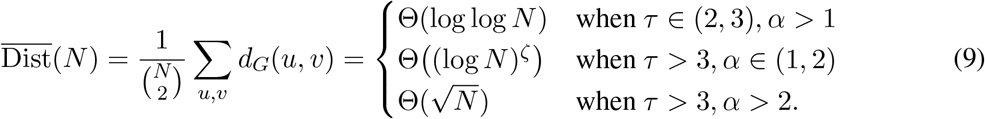

As [21] has found, these average distances give a good indication as to the speed at which epidemics can spread on a network, as well as an indication of the shape of the epidemic curve [21].

Equation (9) demonstrates that increasing *α* decreases the number of long-range edges when *τ >* 3. To see this, note that when *τ >* 3, the node fitnesses *w*_*u*_ (which satisfy the same power law as (8)) are typically less than 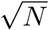 so the ratio

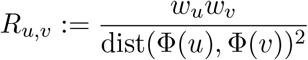

is typically less than 1. In this case, since *R*_*u,v*_ is raise to the power *α* in the connectivity probability (Equation (7)), increasing *α* decreases the probabilities of long-range edges. However, when *τ* ∈ (2, 3), node pairs with ratio *R*_*u,v*_ *>* 1 happen frequently enough, and for these pairs the connection probability is unaffected by the change of *α*. So, networks with *τ* ∈ (2, 3) has many long-range edges irrespective of *α*.

Comparing (9) to the average distance in ageometric network models such as the configuration model (see (10) below) and to the polynomial average distance in lattice models, we observe that geometry and long-range connections play a role in GIRGs with *τ >* 3, and the GIRG model interpolates between the small-world configuration model and the lattice.

Clustering in a network corresponds to the phenomenon known as “a friend of a friend is also likely to be my friend”, and it manifests in the presence of *triangles* in the network. Communities and clustering are naturally present in GIRGs, because the model favours connections between nodes that are close to each other in space. Thus, a GIRG incorporates all five desired features.

For the spread of information or infections in GIRGs and similar networks, it is known that large-degree nodes and many triangles have opposing effects. On the one hand, nodes of large degree (also called hubs, super-spreaders, or influencers) contribute to fast dissemination, and foster explosive propagation of information or infections [50, 51, 52, 53]. On the other hand, clustering and community structures provide natural barriers that slow down the process, while long-range edges accelerate the spread [54, 55, 56, 57, 58]. (In other settings increased clustering may have the opposite effect [60], and sometimes in similar settings the presence of superspreaders may dominate to the point at which clustering has a negligible effect [61].)

### 3.2 The configuration model and other ageometric network models

Ageometric random network models are often used as null-models to compare network data to purely random networks. Here we describe a commonly used model, the configuration model This is a well-studied random-graph model, dating back to Bender and Canfield [59], Bollobás [62] and Wormald [63], and introduced in network science by Molloy and Reed [64, 65]. The main advantage of this simple model is that it can mimic the degree distribution of real-life networks.

#### Definition 5

(Configuration model). *Fix N* ≥ 1 *the number of nodes. Prescribe to each node u* ∈ {1, 2, …, *n*} *its node-degree* deg(*u*) ≥ 0, *so that the total degree is even. h*_*N*_ := Σ_*u* ≤ *N*_ deg(*u*) *To form a graph, to each node u we assign* deg(*u*) *half-edges. The half-edges are then paired uniformly at random to form edges. Self-loops and multiple edges are then erased from the resulting multi-graph*.

It is known that the erasure of self-loops and multiple edges does not affect the empirical degree distribution of the model in any significant way [66].

In comparison to GIRGs, the configuration model can incorporate the desired properties (a), (c) but not (b), (d) and (e). For (a), highly variable degree distributions can be incorporated since the degree sequence is prescribed.

Small distances are an important feature of ageometric network models in general, aka the *small-world property*. Heuristically, this means that two arbitrary nodes in the network can be connected via very short paths, using only a few connections. Mathematically, when setting power law degree distribution, the average distance in the model scales as [67, 68]

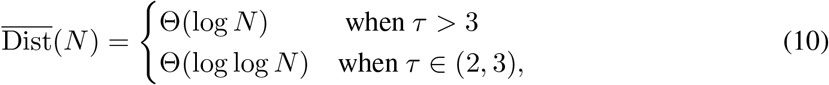

The second row, showing doubly-logarithmic distances, is often called ultra-small world. When *τ >* 3, the shape of the epidemic curve is shown to converge in the large network limit, see [69, 70, 71].

While the configuration model and other ageometric models easily accommodate power-law node degrees, and have the small-world property, they neither contain communities nor clustering [47, 40].

Other popular ageometric models include the Chung-Lu and the similar Norros-Reittu model [72, 73, 74], and preferential attachment models (also known as the Barabási-Albert model) [75]. In the Chung-Lu and the Norros-Reittu model, only the expected degrees of nodes are prescribed, rather than their exact degree, while in preferential attachment models, the power law exponent can be tuned. Using similar parameter settings, these models behave qualitatively similarly to the configuration model, although numerical values might differ, see [66, 76, 47, 77, 78] for references. As a result, we choose the configuration model for baseline comparison.

In [21] we have compared the behaviour of a similar epidemic on the configuration model and on GIRGs. There, we found the epidemic curves are similar for models with the same degree distribution whenever this distribution has a power law with some *τ* ∈ (2, 3). Furthermore, GIRGs with any parameter *τ >* 3 and long-range parameter *α* ∈ (1, 2) are comparable with a configuration model with similar degree distribution. This phenomenon is best explained by comparing the average distance, i.e., (9) to (10): when *τ* ∈ (2, 3), both models have doubly-logarithmic distances and thus allow for very quick spread, while when *τ >* 3 and *α* ∈ (1, 2), poly-logarithmic distances in GIRGs are comparable to logarithmic distances in the ageometric counterpart, hence the similarity of the curves.

While we expect the same universality to occur for the more complicated SEISeR model, for baseline comparison, we also carry out some of our simulations on the configuration model with power law degree distributions, as in (8).

## 4 Supplementary figures

### CRediT Author Contributions

LG contributed to Conceptualization, Investigation, Writing — Original Draft Preparation, Writing — Review & Editing; JJ contributed to Conceptualization, Data Curation, Investigation, Software, Visualisation; Writing — Review & Editing; JK contributed to Conceptualization, Investigation, Writing — Original Draft Preparation, Writing — Review & Editing; JL contributed to Investigation, Software, Validation, Writing — Review & Editing.

## Data Availability

The software can be obtained from the authors. The data is available via 10.5281/zenodo.4601163.

https://doi.org/10.5281/zenodo.4601163

## Acknowledgements

The work of JJ and JK is partly supported by the Netherlands Organisation for Scientific Research (NWO) through grant NWO 613.009.122. There was no additional external funding received for this study.

